# Selection for transcatheter versus surgical aortic valve replacement and mid-term survival: results of the AUTHEARTVISIT Study

**DOI:** 10.1101/2024.02.07.24302456

**Authors:** Johann Auer, Pavla Krotka, Berthold Reichardt, Denise Traxler, Ralph Wendt, Michael Mildner, Hendrik Jan Ankersmit, Alexandra Graf

## Abstract

Limited data are available from randomized trials comparing outcomes between transcatheter aortic valve replacement (TAVR) and surgery in patients with different risks and with follow-up >2 years. In this large, population-based cohort study, long-term mortality and morbidity were investigated in patients undergoing aortic valve replacement (AVR) for severe aortic stenosis using a surgically implanted bioprosthesis (sB-AVR) or TAVR.

Individual data from the Austrian Insurance funds from 2010 through 2020 were analysed. The primary outcome was all-cause mortality, assessed in the overall and propensity score–matched populations. Secondary outcomes included reoperation and cardiovascular events.

From January 2010 through December 2020, a total of 18 882 patients underwent sB-AVR (n=11 749; 62.2%) or TAVR (n=7133; 37.8%); median follow-up was 4.0 (interquartile range 2.1–6.5) years (maximum 12.3 years). The risk of all-cause mortality was higher with TAVR compared with sB-AVR: hazard ratio (HR) 1.552, 95% confidence interval (CI) 1.469–1.640, p<0.001; propensity score–matched HR 1.510, 1.403–1.625, p<0.001. Estimated median survival was 8.8 years (95% CI 8.6– 9.1) with sB-AVR vs 5 years (4.9–5.2) with TAVR. Estimated 5-year survival probability was 0.664 (0.664–0.686) with sB-AVR vs 0.409 (0.378–0.444) with TAVR overall, and 0.690 (0.674–0.707) and 0.560 (0.540–0.582), respectively, with propensity score matching. Other predictors of mortality were age, sex, previous heart failure, diabetes, and chronic kidney disease.

In >2-year follow-up, selection for TAVR was significantly associated with higher all-cause mortality compared with sB-AVR in patients ≥65 years with severe, symptomatic aortic stenosis.

Graphical Abstract

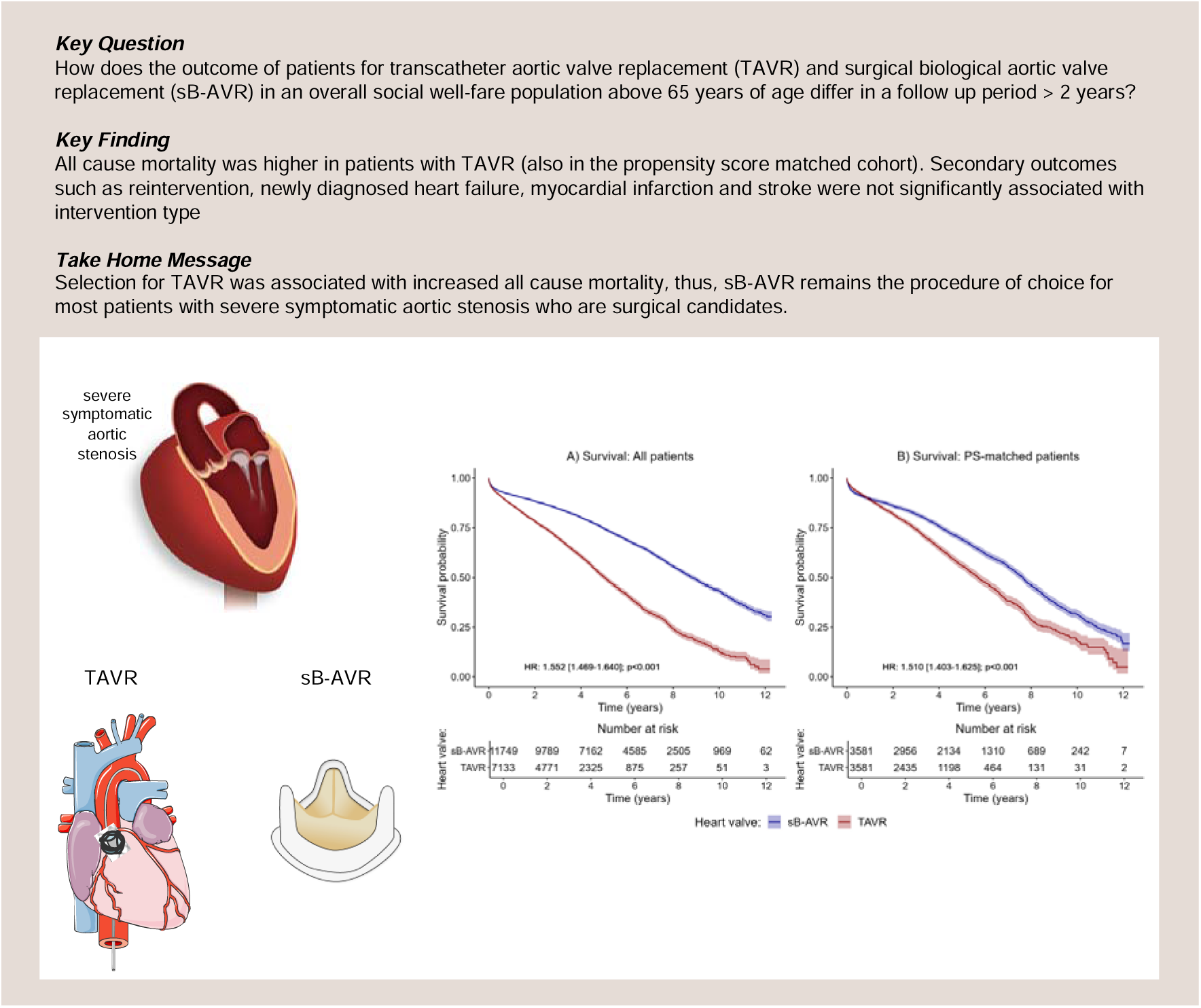

## Introduction

The comparative effectiveness of aortic valve replacement (AVR) for severe aortic stenosis using a surgically implanted bioprosthesis (sB-AVR) or transcatheter aortic valve replacement (TAVR) can vary depending on several factors.[1] In general, surgical AVR (SAVR) is considered to be the gold standard for AVR because it provides a longer-lasting and more durable solution compared with TAVR. However, SAVR requires open-heart surgery, which is associated with a longer recovery time and higher risks of complications compared with minimally invasive TAVR. Both SAVR and TAVR have advantages and disadvantages, and factors involved in procedure selection include patient characteristics, type of valve, and technical aspects of the procedure. TAVR and SAVR differ in length of hospital stay and overall recovery time, paravalvular regurgitation, hemodynamics, ease of coronary access, structural valve deterioration, and need for a new pacemaker. Rates of vascular complications, pacemaker implantations, and paravalvular regurgitation are higher after TAVR. In contrast, severe bleeding events, acute kidney injury, and new-onset atrial fibrillation are more frequent after SAVR.[2]

TAVR has emerged as the most frequently used therapy for older patients with severe symptomatic aortic stenosis and has surpassed SAVR in procedural volume in patients age 75 years or older in the United States and Europe.[3] Randomized controlled trials (RCTs) have demonstrated equivalent or even superior outcomes with TAVR compared with SAVR in elderly patients across a variety of risk categories.[4] RCTs are usually the accepted standard for establishing the efficacy of medical interventions,[1] but they cannot capture every aspect of a therapeutic intervention’s effects in clinical practice.[2, 3] A systematic review and meta-analysis, for example, showed that RCTs comparing TAVR with SAVR had systematic imbalances in the proportion of deviations from random assigned treatment, loss to follow-up, and receipt of additional procedures, including coronary artery bypass surgery. These factors can undermine internal validity because of the related high risk of attrition and performance biases.[5] Real-world evidence is derived from studies conducted with non-randomized data, including data collected by the health care system, such as longitudinal insurance claims.[6] Such evidence can complement clinical trial findings, facilitating a more complete understanding of the effectiveness of therapeutic interventions.

The current analysis is based on data from the AUTHEARTVISIT registry for patients with severe aortic stenosis who had an indication for AVR and were selected to undergo either TAVR or sB-AVR. The results characterize the mid-term and long-term clinical outcomes for this patient group, followed for longer than 2 years.

## Methods

### Study design and patients

This national population-based cohort study complied with the Declaration of Helsinki and was approved by the ethics committee of lower Austria (GS1-EK-4/722-2021). The trial was registered with clincialtrials.gov (NCT05912660). Study data were generated retrospectively by retrieval from the Austrian Health Insurance Fund. Data on outcomes and potential confounding factors were derived from billing information based on MEL (i.e., Medizinische Einzelleistung, or individual medical procedure) and International Classification of Diseases (ICD) codes available for each patient from one year before surgery up to study cut-off. Health care in Austria is a national system with good access to care, few malpractice lawsuits, and little tendency toward overuse of medical resources. Approximately 98% of the Austrian population is registered in the public health insurance system, but a minority of patients paying for medical supplies using private insurance[7] were not included in the AUTHEARTVISIT trial.

This analysis included all patients age ≥65 years registered in the Austrian Health System who underwent AVR with either sB-AVR or TAVR and without coronary revascularization within 4 months before the index procedure in Austria from 1 January 2010 through 31 December 2020. Patients who underwent AVR with a mechanical valve, had concomitant heart surgery, or underwent percutaneous coronary stenting within 4 months prior to the index operation were excluded.

### Study outcomes

The primary study outcome was all-cause mortality. Secondary outcomes were re-operation, stroke, myocardial infarction, and heart failure. For each patient, billing information (based on MEL codes) and diagnoses (based on ICD codes) were available from one year before the index procedure up to study cut-off. Furthermore, death dates were available at the time of study cut-off. To evaluate secondary outcomes (re-operation, stroke, myocardial infarction, heart failure) for each patient, data were scanned from the index procedure through the study cut-off date for the corresponding ICD or MEL codes. A detailed list of all ICD codes used for the outcome variables is shown in the supplementary file (Supplementary Tables 1 and 2).

### Statistical analysis

Continuous data are presented as means ± standard deviation (SD) or as medians with interquartile ranges (IQRs). Categorical data are presented as absolute numbers and percentages. Continuous variables were compared between study groups using Student’s t-test, and categorical variables were compared using the chi-squared test.

To evaluate the association between intervention (sB-AVR or TAVR) and the primary outcome of all-cause mortality, Cox regression was used. To account for potential confounding variables, multivariable Cox regression analysis was performed, with the following variables included: age (per one year increase), sex (male, female), heart failure, myocardial infarction, stroke, diabetes mellitus, adiposity hyperlipidemia, hyperuricemia/gout, valvular/arrhythmogenic/other cardiomyopathies (CMPs), ischemic CMP, atherosclerosis, pulmonary diseases, kidney diseases, or malignant diseases prior to the index operation. These comorbidities were defined using ICD codes available for each patient up to one year before the index procedure. Data available before the index procedure were scanned for each patient based on ICD codes and categorized for the different comorbidities. If an ICD code for a comorbidity as a main or secondary diagnosis was observed at least one time in the year before the index procedure, the patient was assumed to have the comorbidity. Detailed information on ICD codes used for this analysis is shown in the supplementary file (Supplementary Tables 3–5). Schönfeld residuals were used to evaluate the proportional hazard assumption and variance inflation factors to evaluate multicollinearity. Hazard ratios (HRs) are presented as TAVR vs. sB-AVR, so that a HR >1 indicates increased risk of the corresponding event in the TAVR group (and vice versa).

Furthermore, propensity score matching (PSM) was performed for the two study groups (sB-AVR and TAVR). Propensity scores were estimated with logistic regression based on age, sex, heart failure, myocardial infarction, stroke, diabetes mellitus, adiposity, hyperlipidemia, hyperuricemia/gout, valvular/arrhythmogenic/other CMPs, ischemic CMP, atherosclerosis, pulmonary diseases, kidney diseases, and malignant diseases prior to the operation. Matching was performed using the nearest neighbor method (1:1 ratio with a caliper width of 0.0001 or 0.001 depending on the heterogeneity of the investigated subgroup).

To investigate results on all-cause mortality in more detail, several sensitivity analyses were performed. Given indications of a non-proportional hazard behavior for the group variable (crossing survival curves in the PSM groups), we additionally investigated a potential differential effect of group on all-cause death in different time intervals (years 1, 2, 3, 4, and >4) using time-dependent coefficients for the study group. Furthermore, the primary Cox regression analyses for all-cause mortality were repeated for several subgroups of patients: ages 65–75 years, age >75 years, with diabetes mellitus, with pulmonary diseases, with kidney disease, with the index procedure before 2015, and with index procedure during and after 2015. Furthermore, patients surviving the first month after the index procedure were assessed for risk of all-cause mortality based on implantation or not of a pacemaker within that first month.

For the secondary outcomes (reoperation, myocardial infarction, heart failure, stroke), competing risk analyses were performed, using death as a competing risk. Confounding variables for multivariable analysis were the same as those input into the Cox regression model for the primary outcome. In case of heart failure, the analysis was adapted, as patients with a prior diagnosis of heart failure cannot be newly diagnosed and were therefore excluded.

All tests were performed using a two-sided significance level of 0.05. Because of the retrospective and exploratory character of the study, there was no correction for multiple comparisons. Statistical analyses were conducted and graphs generated in R (version 4.1.3) using the following packages: ggplot2 (version 3.4.2), survival (version 3.2.13), survminer (version 0.4.9), cmprsk (version 2.2.11), and MatchIt (version 4.4.0).

## Data availability

Pseudonymized participant data will be made available upon qualified request starting with publication. Approval of a proposal and a signed data access agreement are mandatory.

## Results

### Study population and patient characteristics

The study included 18 882 patients registered in the Austrian Health Funds who underwent AVR with sB-AVR or TAVR from 1 January 2010 through 31 December 2020. sB-AVR was performed in 11 749 (62.2%) and TAVR in 7133 (37.8%). In 2010, sB-AVR was performed in most patients, but over time, TAVR became the more common choice, and in 2019, more patients received TAVR than sB-AVR (Figure 1). We also divided the cohort into patients ages 65–75 years (n=7575, 40.1%) and those age >75 years (n=11 307, 59.9%). In the younger group, sB-AVR was predominant (n=6483, 85.6% vs n=1092, 14.4% receiving TAVR), but not in the older group (n=5266, 46.6% receiving sB-AVR vs n=6041, 53.4%, receiving TAVR). In the younger patient group, sB-AVR was the more common method throughout the follow-up period, in contrast to the older group, in which TAVR became more common in 2015 (Figure 1).

**Figure 1.**
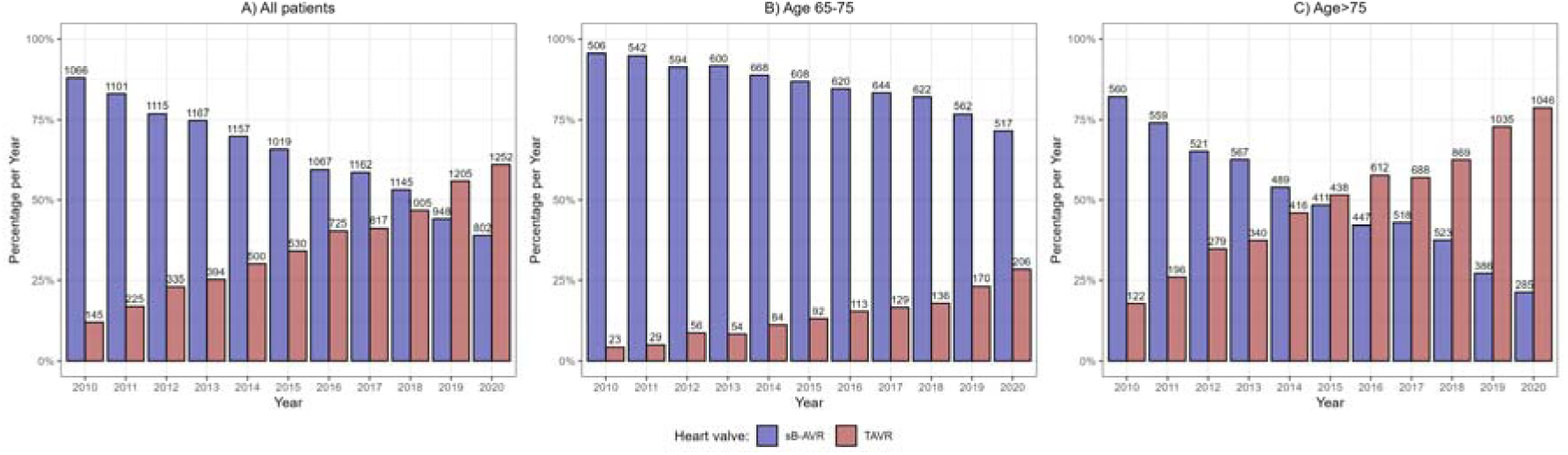
Percentages per year (y-axis) and absolute numbers (above the bars) of patients receiving TAVI or sB-AVR per calendar year for (A) the overall cohort, (B) those ages 65–75 years, and (C) those older than 75 years.

### All-cause mortality

The primary endpoint was all-cause mortality. Kaplan–Meier curves for all patients (Figure 2A) and PSM patients (Figure 2B) showed a significantly increased all-cause mortality with TAVR compared to sB-AVR. Table 1 shows baseline characteristics between both groups before and after PSM. Also standard Cox regression, accounting for several confounding factors, showed an increased all-cause mortality with TAVR (1.552, 95% CI 1.469–1.640, p<0.001) overall and in the PSM analysis (1.510, 1.403–1.625, p<0.001).

**Figure 2.**
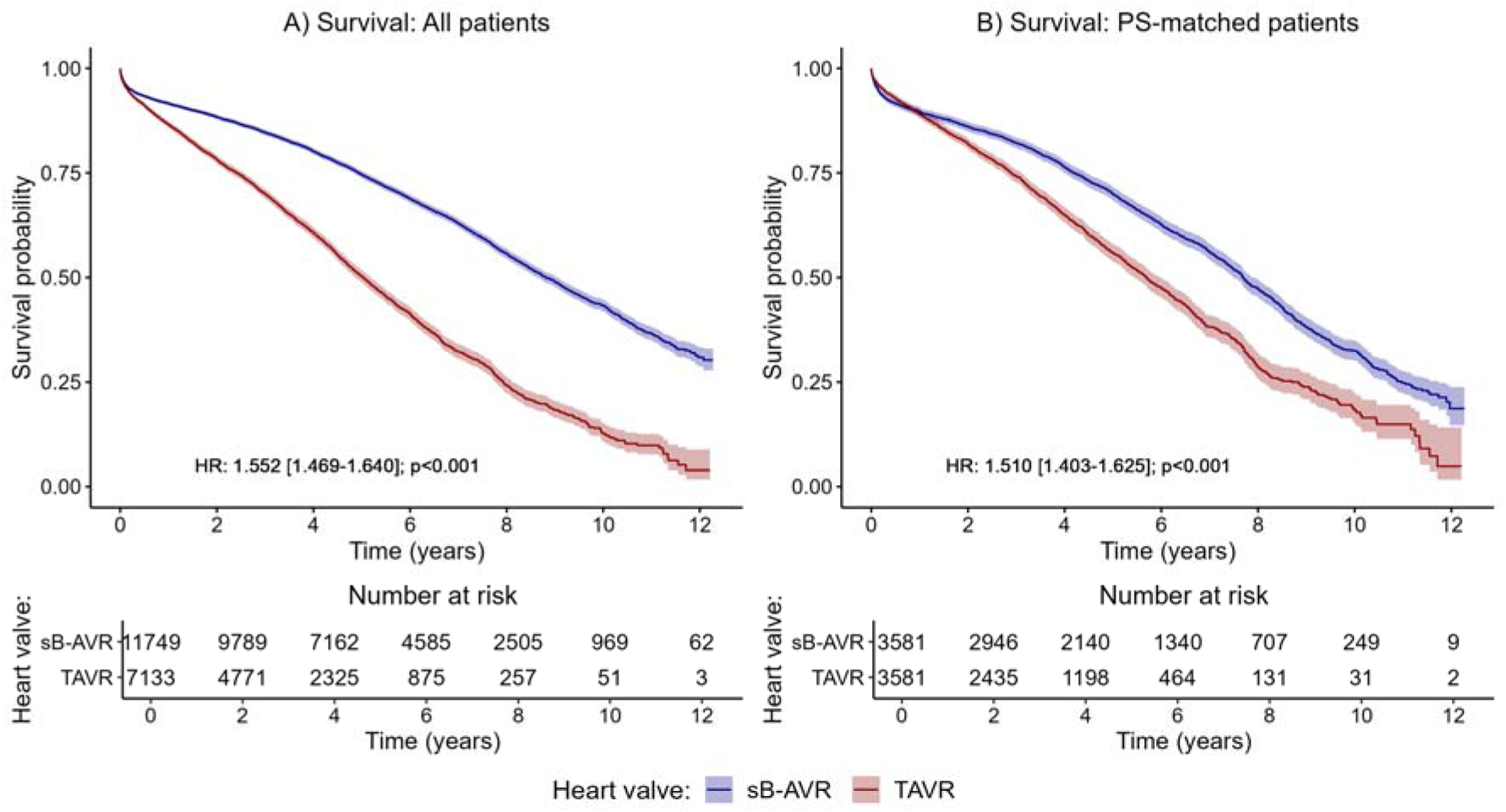
Kaplan–Meier curves and 95% Confidence Intervals for overall survival in (A) all patients and (B) the propensity score–matched groups.

**Table 1.**
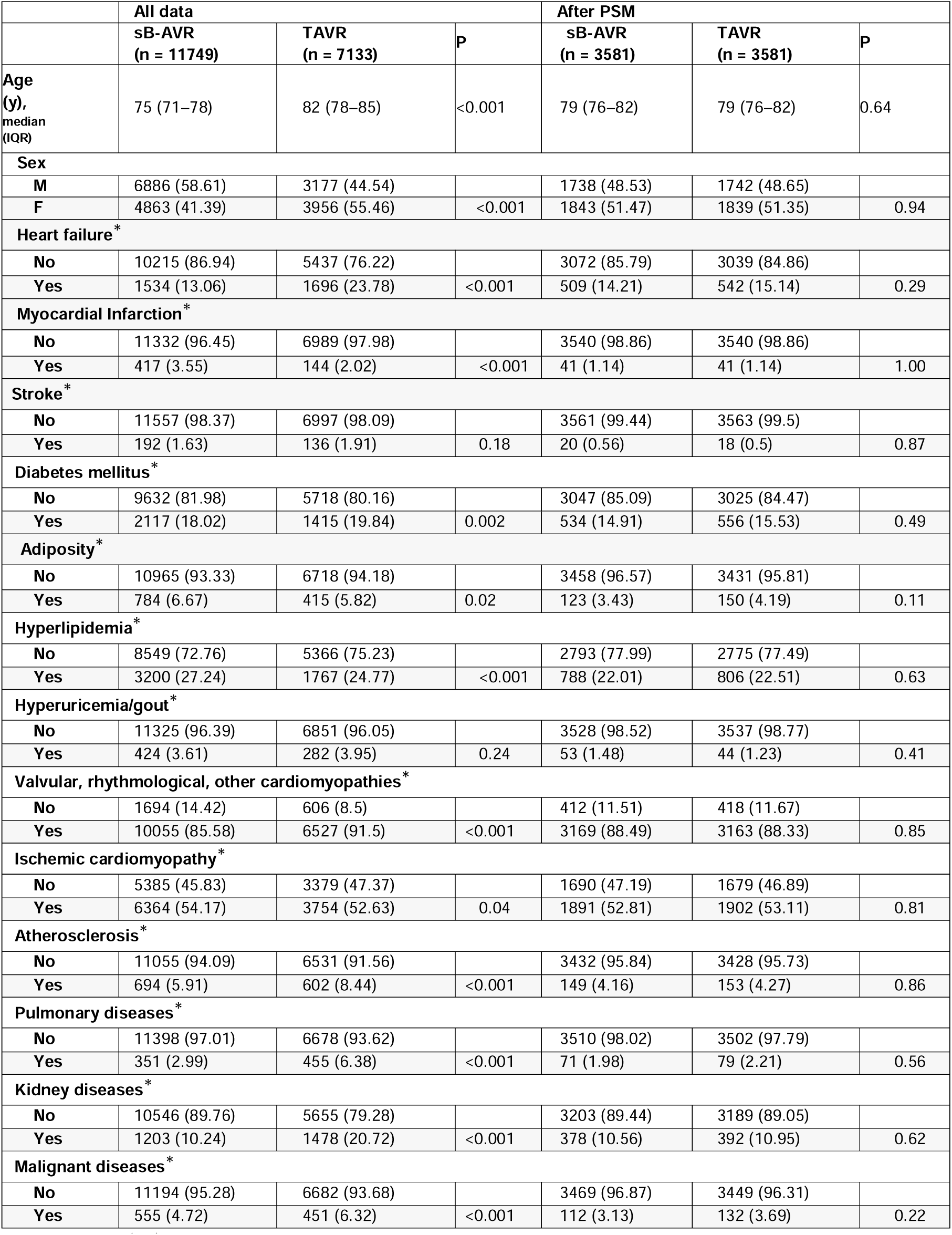
Baseline characteristics compared between the sB-AVR and TAVR groups before and after propensity score matching (PSM); *present before operation.

|  | All data |  |  | After PSM |  |  |
| --- | --- | --- | --- | --- | --- | --- |
|  | sB-AVR<br>(n = 11749) | TAVR<br>(n = 7133) | P | sB-AVR<br>(n = 3581) | TAVR<br>(n = 3581) | P |
| Age<br>(y),<br>median<br>(IQR) | 75 (71–78) | 82 (78–85) | <0.001 | 79 (76–82) | 79 (76–82) | 0.64 |
| **Sex** |  |  |  |  |  |  |
| M | 6886 (58.61) | 3177 (44.54) |  | 1738 (48.53) | 1742 (48.65) |  |
| F | 4863 (41.39) | 3956 (55.46) | <0.001 | 1843 (51.47) | 1839 (51.35) | 0.94 |
| **Heart failure\*** |  |  |  |  |  |  |
| No | 10215 (86.94) | 5437 (76.22) |  | 3072 (85.79) | 3039 (84.86) |  |
| Yes | 1534 (13.06) | 1696 (23.78) | <0.001 | 509 (14.21) | 542 (15.14) | 0.29 |
| **Myocardial Infarction\*** |  |  |  |  |  |  |
| No | 11332 (96.45) | 6989 (97.98) |  | 3540 (98.86) | 3540 (98.86) |  |
| Yes | 417 (3.55) | 144 (2.02) | <0.001 | 41 (1.14) | 41 (1.14) | 1.00 |
| **Stroke\*** |  |  |  |  |  |  |
| No | 11557 (98.37) | 6997 (98.09) |  | 3561 (99.44) | 3563 (99.5) |  |
| Yes | 192 (1.63) | 136 (1.91) | 0.18 | 20 (0.56) | 18 (0.5) | 0.87 |
| **Diabetes mellitus\*** |  |  |  |  |  |  |
| No | 9632 (81.98) | 5718 (80.16) |  | 3047 (85.09) | 3025 (84.47) |  |
| Yes | 2117 (18.02) | 1415 (19.84) | 0.002 | 534 (14.91) | 556 (15.53) | 0.49 |
| **Adiposity\*** |  |  |  |  |  |  |
| No | 10965 (93.33) | 6718 (94.18) |  | 3458 (96.57) | 3431 (95.81) |  |
| Yes | 784 (6.67) | 415 (5.82) | 0.02 | 123 (3.43) | 150 (4.19) | 0.11 |
| **Hyperlipidemia\*** |  |  |  |  |  |  |
| No | 8549 (72.76) | 5366 (75.23) |  | 2793 (77.99) | 2775 (77.49) |  |
| Yes | 3200 (27.24) | 1767 (24.77) | <0.001 | 788 (22.01) | 806 (22.51) | 0.63 |
| **Hyperuricemia/gout\*** |  |  |  |  |  |  |
| No | 11325 (96.39) | 6851 (96.05) |  | 3528 (98.52) | 3537 (98.77) |  |
| Yes | 424 (3.61) | 282 (3.95) | 0.24 | 53 (1.48) | 44 (1.23) | 0.41 |
| **Valvular, rhythmological, other cardiomyopathies\*** |  |  |  |  |  |  |
| No | 1694 (14.42) | 606 (8.5) |  | 412 (11.51) | 418 (11.67) |  |
| Yes | 10055 (85.58) | 6527 (91.5) | <0.001 | 3169 (88.49) | 3163 (88.33) | 0.85 |
| **Ischemic cardiomyopathy\*** |  |  |  |  |  |  |
| No | 5385 (45.83) | 3379 (47.37) |  | 1690 (47.19) | 1679 (46.89) |  |
| Yes | 6364 (54.17) | 3754 (52.63) | 0.04 | 1891 (52.81) | 1902 (53.11) | 0.81 |
| **Atherosclerosis\*** |  |  |  |  |  |  |
| No | 11055 (94.09) | 6531 (91.56) |  | 3432 (95.84) | 3428 (95.73) |  |
| Yes | 694 (5.91) | 602 (8.44) | <0.001 | 149 (4.16) | 153 (4.27) | 0.86 |
| **Pulmonary diseases\*** |  |  |  |  |  |  |
| No | 11398 (97.01) | 6678 (93.62) |  | 3510 (98.02) | 3502 (97.79) |  |
| Yes | 351 (2.99) | 455 (6.38) | <0.001 | 71 (1.98) | 79 (2.21) | 0.56 |
| **Kidney diseases\*** |  |  |  |  |  |  |
| No | 10546 (89.76) | 5655 (79.28) |  | 3203 (89.44) | 3189 (89.05) |  |
| Yes | 1203 (10.24) | 1478 (20.72) | <0.001 | 378 (10.56) | 392 (10.95) | 0.62 |
| **Malignant diseases\*** |  |  |  |  |  |  |
| No | 11194 (95.28) | 6682 (93.68) |  | 3469 (96.87) | 3449 (96.31) |  |
| Yes | 555 (4.72) | 451 (6.32) | <0.001 | 112 (3.13) | 132 (3.69) | 0.22 |

The choice of valve type was only one factor in survival, and most of the other included variables were significantly associated with all-cause mortality in the overall and PSM analyses (Table 2, standard Cox model). Median follow-up was 4.02 (IQR 2.1–6.5) years in the overall population, 2.9 (1.6–4.9) years with TAVR, and 5.0 (2.8– 7.6) years in the sB-AVR group. Estimated median survival time was 8.8 years (95% CI 8.6–9.1) in the sB-AVR group, compared with 5 years (4.9–5.2) in the TAVR group. In more detail, the estimated 5-year survival probability was 0.664 (95% CI 0.664–0.686) in the sB-AVR group, compared with 0.409 (0.378–0.444) in the TAVR group. PSM changed this estimate to 0.690 (0.674–0.707) with sB-AVR vs 0.560 (0.540–0.582) with TAVR.

**Table 2.** Hazard ratios (HRs) and corresponding 95% confidence intervals (CIs) from Cox regressions for all-cause mortality in all patients and the propensity score– matched patients for the standard and time-dependent models; *present before operation

|  | Standard Cox model |  |  |  | Time-dependent analyses |  |  |  |
| --- | --- | --- | --- | --- | --- | --- | --- | --- |
|  | All patients |  | Propensity score-matched |  | All patients |  | Propensity score-matched |  |
|  | HR (95% CI) | P | HR (95% CI) | p-value | HR (95% CI) | P | HR (95% CI) | P |
| Heart valve (sB-AVR vs. TAVR) | 1.552<br>(1.469–1.640) | **<0.001** | 1.510<br>(1.403–1.625) | **<0.001** |  |  |  |  |
| Year 1 |  |  |  |  | 1.073<br>(0.977–1.178) | **0.14** | 1.044<br>(0.905–1.203) | **0.56** |
| Year 2 |  |  |  |  | 1.902<br>(1.662–2.176) | **<0.001** | 2.006<br>(1.615–2.492) | **<0.001** |
| Year 3 |  |  |  |  | 1.746<br>(1.520–2.005) | **<0.001** | 1.971<br>(1.589–2.445) | **<0.001** |
| Year 4 |  |  |  |  | 1.796<br>(1.563–2.064) | **<0.001** | 1.704<br>(1.393–2.083) | **<0.001** |
| Year >5 |  |  |  |  | 1.750<br>(1.611–1.901) | **<0.001** | 1.574<br>(1.397–1.774) | **<0.001** |
| Age (per one year increase) | 1.053<br>(1.049–1.058) | **<0.001** | 1.036<br>(1.028–1.044) | **<0.001** | 1.053<br>(1.049–1.058) | **<0.001** | 1.036<br>(1.028–1.044) | **<0.001** |
| Sex (male vs. female) | 0.865<br>(0.826–0.906) | **<0.001** | 0.872<br>(0.812–0.937) | **<0.001** | 0.863<br>(0.824–0.904) | **<0.001** | 0.872<br>(0.812–0.937) | **<0.001** |
| Heart failure* | 1.420<br>(1.343–1.502) | **<0.001** | 1.373<br>(1.248–1.511) | **<0.001** | 1.424<br>(1.346–1.506) | **<0.001** | 1.375<br>(1.250–1.513) | **<0.001** |
| Myocardial Infarction* | 1.166<br>(1.032–1.317) | **0.01** | 1.427<br>(1.075–1.894) | **0.01** | 1.158<br>(1.025–1.308) | **0.02** | 1.425<br>(1.074–1.891) | **0.01** |
| Stroke* | 1.214<br>(1.031–1.428) | **0.02** | 0.952<br>(0.573–1.583) | **0.85** | 1.210<br>(1.029–1.424) | **0.02** | 0.951<br>(0.572–1.581) | **0.85** |
| Diabetes mellitus* | 1.379<br>(1.303–1.460) | **<0.001** | 1.299<br>(1.177–1.434) | **<0.001** | 1.381<br>(1.305–1.461) | **<0.001** | 1.299<br>(1.177–1.434) | **<0.001** |
| Adiposity* | 1.118<br>(1.021–1.225) | **0.02** | 1.118<br>(0.936–1.336) | **0.22** | 1.116<br>(1.019–1.222) | **0.02** | 1.118<br>(0.936–1.335) | **0.22** |
| Hyperlipidemia* | 0.825<br>(0.781–0.871) | **<0.001** | 0.822<br>(0.750–0.900) | **<0.001** | 0.824<br>(0.781–0.871) | **<0.001** | 0.822<br>(0.750–0.900) | **<0.001** |
| Hyperuricemia/gout* | 1.089<br>(0.972–1.219) | 0.14 | 1.121<br>(0.841–1.496) | **0.44** | 1.091<br>(0.974–1.222) | **0.13** | 1.120<br>(0.839–1.493) | **0.44** |
| Valvular, rhythmological, other cardiomyopathies* | 0.865<br>(0.804–0.931) | **<0.001** | 0.837<br>(0.748–0.937) | **0.002** | 0.866<br>(0.804–0.932) | **<0.001** | 0.838<br>(0.748–0.938) | **0.002** |
| Ischemic cardiomyopathy* | 1.079<br>(1.029–1.132) | **0.002** | 1.122<br>(1.040–1.209) | **0.003** | 1.079<br>(1.029–1.132) | **0.002** | 1.121<br>(1.040–1.208) | **0.003** |
| Atherosclerosis* | 1.205<br>(1.109–1.308) | **<0.001** | 1.229<br>(1.043–1.448) | **0.01** | 1.204<br>(1.109–1.307) | **<0.001** | 1.227<br>(1.042–1.446) | **0.01** |
| Pulmonary diseases* | 1.310<br>(1.167–1.470) | **<0.001** | 1.263<br>(0.970–1.645) | **0.08** | 1.330<br>(1.186–1.493) | **<0.001** | 1.267<br>(0.972–1.650) | **0.08** |
| Kidney diseases* | 1.406<br>(1.322–1.496) | **<0.001** | 1.508<br>(1.352–1.681) | **<0.001** | 1.410<br>(1.325–1.500) | **<0.001** | 1.508<br>(1.353–1.682) | **<0.001** |
| Malignant diseases* | 1.424<br>(1.306–<br>1.553) | **<0.001** | 1.354<br>(1.136–<br>1.614) | **<0.001** | 1.428<br>(1.310–<br>1.557) | **<0.001** | 1.359<br>(1.140–<br>1.620) | **<0.001** |

The results suggested a varying effect of study group on the risk of all-cause mortality over time (crossing survival curves in the PSM patients in the first year after the index procedure; Figure 2B). For this reason, we accounted for a potential time-varying trend using a Cox model with time-dependent coefficients for the grouping factor (Table 2, time-dependent analyses). The analyses of all patients and of PSM patients in the first year after the index procedure showed no significant difference between TAVR and sB-AVR (all patients: HR 1.073, 95% CI 0.977–1.178, p=0.14; PSM patients: 1.044, 0.905–1.203, p=0.56; Table 2). However, for follow-up times >1 year, as already found in the standard Cox model, a significantly higher risk for all-cause mortality was found in patients receiving TAVR, indicating that improved survival with sB-AVR may be evident only with longer follow-up times.

The effect of a significantly higher risk for all-cause mortality in TAVR as compared with sB-AVR was even more prominent in patients ages 65–75 years (Figure 3A, B; HR 2.455, 95% CI 2.209–2.728, p<0.001; Figure 3A). This effect was also observed in the PSM group (HR 2.368, 2.019–2.779, p<0.001; Figure 3B). In the time-dependent analyses, across all separately investigated time intervals, a significantly better survival was found (Supplementary Table 6). Details on baseline data for the subgroup of patients ages 65–75 years and the corresponding PSM patients are given in Supplementary Table 7.

**Figure 3.**
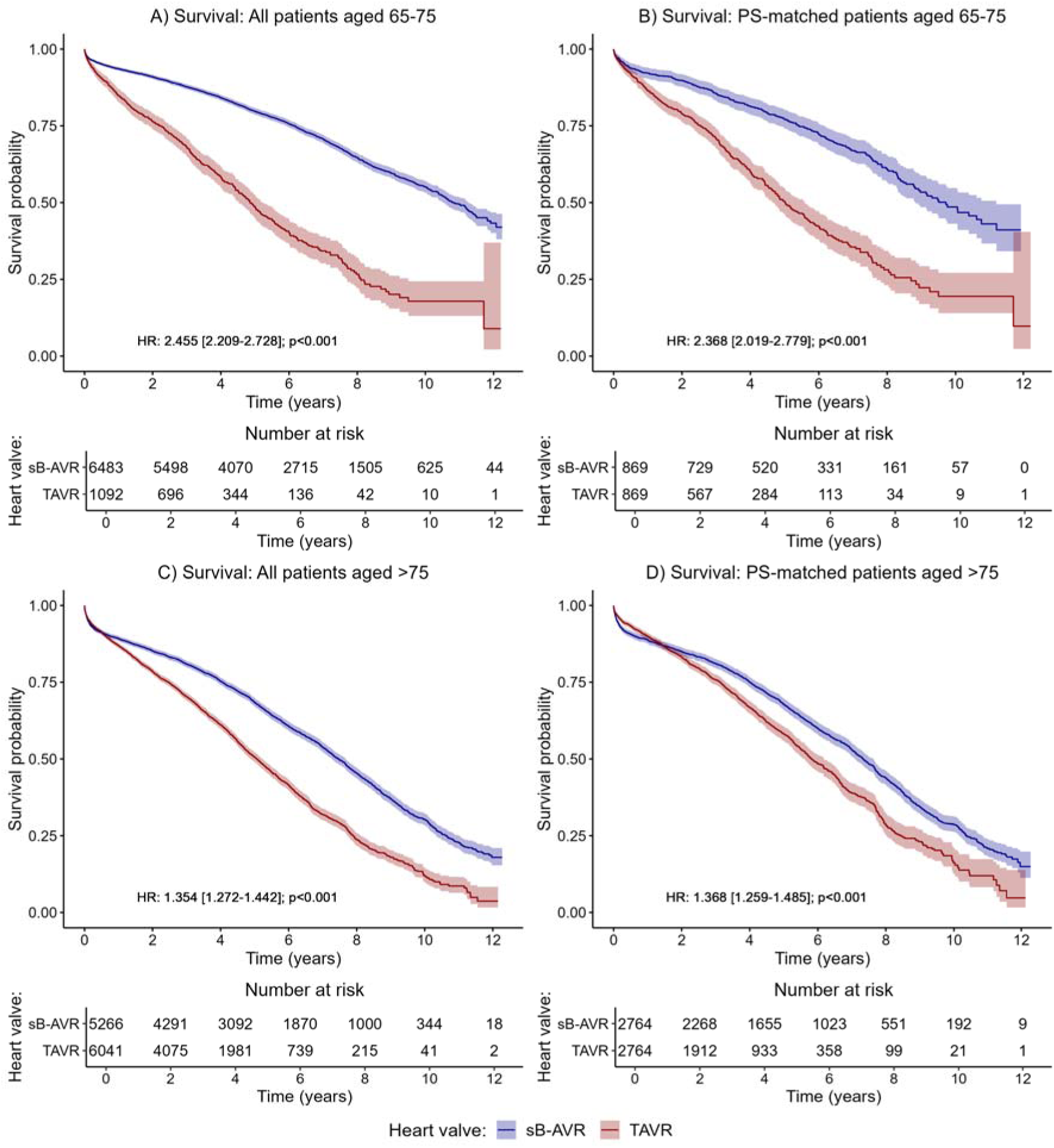
Kaplan–Meier curves and 95% Confidence intervals for overall survival for subgroups of patients ages 65–75 years in the (A) overall cohort and (B) PSM groups and those age >75 years in the (C) overall cohort and (D) PSM groups.

Of note, in older patients (>75 years), TAVR performed significantly worse for the primary outcome of all-cause mortality overall (HR 1.345, 95% CI 1.272–1.442, p<0.001; Figure 3C) and in the PSM patients (1.368, 1.259–1.485, p<0.001; Figure 3D). The time-dependent analyses again showed no significant difference between groups in the first year after the index procedure but a significantly better survival in the sB-AVR group with longer follow-up times (Supplementary Table 8). Details on baseline data for the subgroup of patients >75 years and corresponding PSM patients can be found in Supplementary Table 9.

All multivariable models showed that the choice of AVR is only one factor in a complex system (Table 2, Supplementary Tables 6 and 8). To evaluate the results in more detail, we performed several sensitivity analyses (for detailed results, see Supplementary file, chapters 4 to 6). In separate analyses of all-cause mortality in patients treated before 2015 as well as during and after 2015, TAVR patients still had a significant larger risk for all-cause mortality as compared to sB-AVR in both subsets (before 2015: HR: 1.570, 95% CI 1.456–1.690, p<0.001; during/after 2015: HR: 1.704, 1.562–1.860, p<0.001; Supplementary Figure 1, Supplementary Table 10).

Furthermore, we investigated all-cause mortality by intervention type in three patient subsets defined by underlying disease (diabetes, pulmonary diseases, kidney diseases). In all three subsets, all-cause mortality was significantly increased in patients with TAVR as the primary intervention. Of the 3532 patients with diabetes before the intervention, 1735 died within the follow-up period (sB-AVR, n=988; TAVR, n=747). TAVR as the primary intervention was associated with significantly higher risk for all-cause mortality (HR 1.587, 1.418–1.777, p<0.001; Supplementary Figure 2A, Supplementary Table 11). The subset with pulmonary diseases was notably smaller at 806 patients, of whom 108 in the sB-AVR group and 204 in the TAVR group died. For this patient group, TAVR was significantly associated with survival larger risk for all-cause mortality (HR 2.057, 1.556–2.720, p<0.001; Supplementary Figure 2B, Supplementary Table 11). In the 2681 patients with kidney diseases at the date of the primary intervention, 606 in the sB-AVR group and 813 in the TAVR group died during follow-up (TAVR vs sB-AVR, HR 1.436, 1.269–1.625, p<0.001; Supplementary Figure 2C, Supplementary Table 11).

Pacemaker implantation is common after AVR, so we also analyzed all-cause mortality in patients with implantation within the first month after AVR (Supplementary Figure 3, Supplementary Table 12). A total of 662 patients were excluded from this subanalysis (394 sB-AVR and 268 TAVR) because they died within the first month after the procedure and could not have received a pacemaker. Patients who underwent TAVR had a higher incidence of permanent pacemaker implantation (11.22%) compared with those who had sB-AVR (4.5%). Patients receiving a pacemaker had a significantly higher risk of all-cause death compared with patients who did not receive a pacemaker (HR 1.107, 1.016–1.207, p=0.02).

### Secondary outcomes

Of the 287 patients needing aortic valve reintervention, 232 had undergone sB-AVR and 55 TAVR. Risk of reoperation was not significantly increased by intervention type (HR 0.822, 95% CI 0.597–1.132, p=0.23) in the overall patient cohort (Supplementary Figure 4A, Supplementary Table 13).

As noted, not all patients were included in the analyses for newly diagnosed heart failure. Of the 15 652 patients who were included, 3671 in the overall cohort developed heart failure, 2339 who had sB-AVR and 1332 who had TAVR. The risk of developing heart failure did not differ significantly with valve type (HR 1.030, 0.949– 1.118, p=0.47; Supplementary Figure 4B, Supplementary Table 13).

During the study follow-up, 443 patients had a myocardial infarction, 302 who had sB-AVR and 141 who had TAVR. Valve type did not show a significant association with the risk for myocardial infarction (HR 1.069, 0.837–1.365, p=0.59; Supplementary Figure 4C, Supplementary Table 13).

Stroke occurred in 1594 patients after the index procedure, 1057 who had sB-AVR and 537 who had TAVR. Intervention type was not significantly associated with stroke risk (HR 0.908, 0.803–1.027, p=0.12; Supplementary Figure 4D, Supplementary Table 13).

## Discussion

In this long-term population-based cohort study, undergoing sB-AVR versus TAVR was significantly associated with lower all-cause mortality in patients age 65 years or older with severe, symptomatic aortic stenosis. For a more accurate comparison of outcomes, we used PSM to compare TAVR and sB-AVR patients who were similar with respect to comorbidities[8] and also identified a survival advantage with sB-AVR. Most other candidate factors, including age, sex, previous heart failure, diabetes, and chronic kidney disease, were significantly associated with survival in the overall and PSM cohorts.

These findings are discordant with those of randomized trials in patients with intermediate risk[9] or low risk[10, 11] and with recent meta-analyses.[12, 13] Although randomization allows for control of confounding at baseline, post-randomization exclusion and protocol deviations may still introduce bias.[2, 14] Non-randomized studies using insurance claims databases can be leveraged for real-world evidence, but concerns exist about whether such studies yield unbiased estimates of therapeutic interventions.[15] In RCTs, concomitant procedures (revascularization and others) have not been balanced between patients undergoing TAVR versus sB-AVR, risking performance bias.[16, 17] In the present pragmatic observational study, inclusion of patients with pure symptomatic aortic stenosis without significant coronary artery disease yielded a possibly unique study population for comparing the two treatment strategies without the influence of additional procedures, including revascularization. Divergent results between RCTs and real-world studies that address the same scientific question may be disquieting,[18] but these divergences demonstrate the need to continue testing assumptions that beneficial treatment effects in RCTs will be borne out in practice.[19]

TAVR is associated with lower early complication rates and shorter recovery times, suggesting that it might be an option in high-risk and some elderly patients.[19] Remarkably, our separate survival analyses for patients ages 65–75 years and >75 years indicated a significant survival benefit with sB-AVR in both groups, however, in agreement with randomized trial results suggesting no association between these factors and different mortality risk estimates at long-term follow-up.[4]

We found similar survival with TAVR and sB-AVR up to one year, also in accordance with previous randomized studies of high-risk[20, 21] and intermediate-risk patients with 1–5 years of follow-up.[9, 22, 23] Beyond one year, however, the Kaplan–Meier curves diverged in favor of sB-AVR, including in our PSM patient comparison. Longer post-procedural observation in previous trials might have revealed similar changes depending on patient selection for either AVR strategy.

As with any new technique or technology, practitioners become more efficient with experience, and outcomes might improve.[24] We performed separate analyses of all-cause mortality in patients treated before and during/after 2015 to take potential improvements and technological innovation into account. Results were similar for both observation periods.

Our findings for outcomes with TAVR versus sB-AVR in three subsets of patients with underlying conditions (diabetes, pulmonary diseases, kidney diseases) were consistent with the results for the total study population. Thus, even in patients with these increased risks, long-term outcomes were better in those selected for sB-AVR. In addition, selection for TAVR compared with sB-AVR has consistently been associated with a higher risk of permanent pacemaker implantation. We confirmed this finding, and the higher rate of implantations might have contributed to a higher all-cause mortality rate in patients selected for TAVR.[25]

TAVR could be associated with a higher risk of structural degeneration of the prosthetic valve compared with surgical AVR,[26] risking reoperation and worse outcomes for these patients. We found no significantly higher risk of reoperation with either AVR procedure, however. TAVR also is associated with a specific humoral immune response against alpha-Gal and nonspecific humoral inflammation.[27, 28] A chronic inflammatory response may be associated with increased risk for several conditions that might affect prognosis and overall outcome,[29] but the systemic immune response is reported not to differ after TAVR versus sB-AVR.[30] Any contribution of a differential immunologic response to differential outcomes thus is unlikely.

The main strength of this study is the use of a large, representative real-world national database and allocation of patients to AVR procedures essentially based on contemporary international guidelines. The observational design, however, is a limitation, which we sought to mitigate by including several potential confounding factors in the statistical models and performing detailed PSM to support meaningful comparisons. As in any observational research, even with the large sample size and a long-term follow-up in the current work, unmeasured confounding is still possible. Nevertheless, properly adjusted analyses from large registries can support therapeutic decision-making, and our findings undoubtedly reflect real-world results in patients selected for TAVR or sB-AVR.

Another limitation is that our data did not include all parameters needed for accurate calculation of surgical risk scores, and we could not calculate frailty scores from our database. Despite obtaining data on further diagnoses and medication, we could not eliminate the possibility that sB-AVR was performed more often in healthier patients. Several subgroup analyses did reveal similar effects in differently characterized cohorts.

Our data do not include a proportion of the most recent prosthetic valve devices used for sB-AVR and TAVR, although we do not expect different results with the latest devices. Finally, the data were derived from billing information and discharge coding, requiring an assumption of correct nationwide coding that cannot be retrospectively verified or corrected. Thus, there is the possibility of biased results and conclusions compared with data from prospective clinical databases.

In conclusion, these mid-to long-term population-based findings suggest that selection for sB-AVR compared with TAVR was significantly associated with lower all-cause mortality in patients age 65 years or older with severe, symptomatic aortic stenosis. We did not identify a subgroup of patients who had better outcomes with TAVR, but older patients with high surgical risk may still benefit, at least short term, from a less-invasive approach. With its longer-lasting and more durable resolution compared with TAVR, however, sB-AVR may remain the procedure of choice for severe symptomatic aortic stenosis in patients who are surgical candidates. In this group, sB-AVR was associated with significantly lower mortality rates compared with TAVR.

## Data Availability

All data produced in the present study are available upon reasonable request to the authors

## Acknowledgments

We thank the Pharmaco-economics Advisory Council of the Austrian Sickness Funds for providing the data, especially Ms. Karin Allmer for quality assurance of the database query and Mr. Ludwig Weissengruber for organizational support for data generation.

This work has been presented at the 37^th^ EACTS annual meeting in Vienna.

## Author Contributions

JA and HJA are responsible for conceptualization. PK, BR, AG and HJA conceived the study and curated data. HJA, JA, AG, ML, DT, and PK wrote the paper and visualized the data. PK and AG cleaned, analyzed, and verified the underlying data. HJA provided funding for the paper. All authors commented on the paper, oversaw the analysis, and edited the final manuscript. All authors contributed to the study design. All authors contributed to drafting the paper and revised the manuscript for important intellectual content. All authors had full access to all of the data in the study and had final responsibility for the decision to submit for publication.

