## Supplementary File for "Selection for transcatheter versus surgical aortic valve replacement and mid-term survival: results of the AUTHEARTVISIT Study"

### 1.) Definitions of primary and secondary outcomes

For each patient, billing information (based on MEL codes) and diagnoses (based on ICD codes) were available from one year before the index operation (index-op) to the study cut-off date. Furthermore, death dates were available at the time of study cut-off.

To evaluate the diagnoses, the International Statistical Classification of diseases and Related Health Problems in its 10th revision (ICD-10 version of 2019) was used, which is available on:

<https://icd.who.int/browse10/2019/en>

The corresponding German version is available on:

<https://www.dimdi.de/static/de/klassifikationen/icd/icd-10-who/kode-suche/htmlamtl2019/>

To evaluate outcomes for each patient, data were scanned from index-op to the study cut-off date for the corresponding outcome codes, shown in Supplementary Tables 1 and 2.

#### Table S1. Definitions of primary and secondary outcomes

| **Definitions for Primary and Secondary Outcomes** | |
| --- | --- |
| **Outcome** | **Definition** |
| All-cause Mortality | all-cause mortality based on death date (primary outcome) |
| Reoperation | based on billing information (MEL-Code): first event after index-op with MEL code defined as in Table S2 (secondary outcome) |
| Heart Attack | Based on ICD-Codes: first event after index-op with one of the following ICD-codes: I21.0, I21.1, I21.2, I21.3, I21.4, I21.9 (secondary outcome) |
| Heart Failure | Based on ICD-Codes: first event after index-op with one of the following ICD-codes: I11.0, I13.0, I13.2, I50.0, I50.1, I50.9, I50.11, I50.12, I50.13, I50.14, I50.19 (secondary outcome) |
| Stroke | Based on ICD-Codes: first event after index-op with one of the following ICD-codes: I63.0, I63.1, I63.2, I63.3, I63.4, I63.5, I63.6, I63.8, I63.9, G45.9, G45.0, G45.1, G45.2, G45.3, G45.4, G45.8, I61.0, I61.1, I61.2, I61.3, I61.4, I61.5, I61.6, I61.8, I61.9, I64 (secondary outcome) |

#### Table S2. Definition of outcome “reoperation”

| **Coding for Outcome Reoperation** | |
| --- | --- |
| **MEL Code** | **Description** |
| DB020 | percutaneous implantation of a pulmonary valve |
| DB025 | Aortic valve replacement – catheter directed, transapical, TAVR |
| DB026 | Aortic valve replacement – catheter directed, transvalvular, TAVR |
| DB030 | Reconstruction of the aortic valve |
| DB040 | Reconstruction of the mitral valve |
| DB050 | Reconstruction of the tricuspid valve |
| DB055 | Reconstruction of the pulmonary valve |
| DB060 | Replacement of aortic valve with pulmonary autograft |
| DB070 | Replacement of aortic valve with stentless valve |
| DB080 | Replacement of aortic valve with stented valve |
| DB082 | Replacement of aortic valve with artificial mechanical valve |
| DB090 | Replacement of mitral valve with stentless valve |
| DB100 | Replacement of mitral valve with stented valve |
| DB102 | Replacement of mitral valve with artificial mechanical valve |
| DB110 | Replacement of tricuspid valve with stentless valve |
| DB120 | Replacement of tricuspid valve with stented valve |
| DB122 | Replacement of tricuspid valve with artificial mechanical valve |
| DB130 | Replacement of pulmonary valve with stentless valve |
| DB140 | Replacement of pulmonary valve with stented valve |
| DB142 | Replacement of pulmonary valve with artificial mechanical valve |
| DB021 | Aortic valve replacement – percutaneous, interventional, TAVR |
| XN010 | Aortic valve replacement – percutaneous, interventional, TAVR |

### 2.)  Definitions of confounders and comorbidities

The index-op group was defined using billing information (based on MEL codes); see Table S3.

#### Table S3. Definition of grouping variable (TAVR, sB-AVR)

| **Coding for Group Bio, TAVR** | | |
| --- | --- | --- |
| **MEL Code** | **Description** | **Group Analysis** |
| DB025 | Aortic valve replacement – catheter directed, transapikal, TAVR | TAVR |
| DB026 | Aortic valve replacement – catheter directed, transvalvular, TAVR | TAVR |
| DB060 | Replacement of aortic valve with pulmonary autograft | Biological Valve |
| DB070 | Replacement of aortic valve with stentless valve | Biological Valve |
| DB080 | Replacement of aortic valve with stented valve | Biological Valve |
| DB021 | Aortic valve replacement – percutaneous, interventional, TAVR | TAVR |
| XN010 | Aortic valve replacement – percutaneous, interventional, TAVR | TAVR |

For an additional sensitivity analyses, patients were further divided by receiving or not receiving a pacemaker one month after the index-op. Table S4 shows the MEL codes for pacemaker receipt.

#### Table S4. Definitions for pacemaker receipt

| **Coding for Pacemaker** | |
| --- | --- |
| **MEL Code** | **Description** |
| DE080 | Pacemaker implantation, one chamber system (per session)  pacemaker implantation with one electrode (lead implantation not included) |
| DE081 | Pacemaker implantation, one chamber system, MRI conditional (per session)  pacemaker implantation with one electrode, MRI conditional (lead implantation not included) |
| DE090 | Pacemaker implantation, two chamber system (per session)  pacemaker implantation with atrial and ventricular electrode (lead implantation not included) |
| DE091 | Pacemaker implantation, two chamber system, MRI conditional (per session)  pacemaker implantation with atrial and ventricular electrode, MRI conditional (lead implantation not included) |
| DE100 | Implantation of a cardiac resynchronisation device (per session)  pacemaker implantation with atrial electrode (reatrial), ventricular electrode (reventricular) and left ventricular electrode for av-synchronous biventricular stimulation (artrio-biventricular pacemaker) (probe implantation not included) |
| DE130 | Cardiac pacemaker lead change (per session)  Change of one or multiple leads |
| DE140 | Pacemaker generator change, one chamber system (per session)  Pacemaker generator change without lead change (one lead system) |
| DE141 | Pacemaker generator change, one chamber system, MRI conditional (per session)  Pacemaker generator change without lead change, MRI conditional (one lead system) |
| DE150 | Pacemaker generator change, two chamber system (per session)  Pacemaker generator change without lead change (two lead system) |
| DE151 | Pacemaker generator change, two chamber system, MRI conditional (per session)  Pacemaker generator change without lead change, MRI conditional (two lead system) |

Comorbidities were defined using ICD codes available for each patient up to one year before the index-op. Data available before the index-op were scanned for each patient based on ICD codes categorized for different comorbidities (Table S5). If at least one time in the year before the index-op an ICD code for a comorbidity was observed as a main or secondary diagnosis, the patient was assumed to have the comorbidity.

#### Table S5. ICD codes for comorbidities

| Co-Morbidities | ICD-Codes |
| --- | --- |
| Diabetes mellitus | E10.0, E10.1, E10.2, E10.3, E10.4, E10.5, E10.6, E10.7, E10.8, E10.9, E11.0, E11.1, E11.2, E11.3, E11.4, E11.5, E11.6, E11.7, E11.8, E11.9, E12.0, E12.1, E12.2, E12.3, E12.4, E12.5, E12.6, E12.7, E12.8, E12.9, E13.0, E13.1, E13.2, E13.3, E13.4, E13.5, E13.6, E13.7, E13.8, E13.9, E14.0, E14.1, E14.2, E14.3, E14.4, E14.5, E14.6, E14.7, E14.8, E14.9 |
| Adiposity | E65, E66.0, E66.1, E66.2, E66.8, E66.9 |
| Hyperlipidemia | E78.0, E78.1, E78.2, E78.3, E78.4, E78.5, E78.6, E78.8, E78.9 |
| Hyperuricemia/Gout | E79.0, E79.8, M10.0, M10.00, M10.01, M10.02, M10.03, M10.04, M10.05, M10.06, M10.07, M10.08, M10.09 |
| Valvular, rhythmological and other CMPs | I01.0, I01.1, I01.2, I01.8, I01.9, I02.0, I02.9, I05.0, I05.1, I05.2, I05.8, I05.9, I06.0, I06.1, I06.2, I06.8, I06.9, I07.0, I07.1, I07.2, I07.8, I07.9, I08.0, I08.1, I08.2, I08.3, I08.8, I08.9, I09.0, I09.1, I09.2, I09.8, I09.9, I10, I11.0, I11.9, I12.0, I12.9, I13.0, I13.1, I13.2, I13.9, I15.0, I15.1, I15.2, I15.8, I15.9, I26.0, I26.9, I27.0, I27.1, I27.2, I27.8, I27.9, I28.0, I28.1, I28.8, I28.9, I30.0, I30.1, I30.8, I30.9, I31.0, I31.1, I31.2, I31.3, I31.8, I31.9, I32.0, I32.1, I32.8, I33.0, I33.9, I34.0, I34.1, I34.2, I34.8, I34.9, I35.0, I35.1, I35.2, I35.8, I35.9, I36.0, I36.1, I36.2, I36.8, I36.9, I37.0, I37.1, I37.2, I37.8, I37.9, I38, I39.0, I39.1, I39.2, I39.3, I39.4, I39.8, I40.0, I40.1, I40.8, I40.9, I41.0, I41.1, I41.2, I41.8, I42.0, I42.1, I42.2, I42.3, I42.4, I42.5, I42.6, I42.7, I42.8, I42.9, I43.0, I43.1, I43.2, I43.8, I44.0, I44.1, I44.2, I44.3, I44.4, I44.5, I44.6, I44.7, I45.0, I45.1, I45.2, I45.3, I45.4, I45.5, I45.6, I45.8, I45.9, I46.0, I46.1, I46.9, I470., I47.1, I47.2, I47.9, I48.0, I48.1, I48.2, I48.3, I48.4, I48.9, I49.0, I49.1, I49.2, I49.3, I49.4, I49.5, I49.8, I49.9, I50.0, I50.11, I50.12, I50.13, I50.14, I50.19, I50.9, I51.0, I51.1, I51.2, I51.3, I51.4, I51.5, I51.6, I51.7, I51.8, I51.9, I52.0, I52.1, I52.8, Q20.0, Q20.1, Q20.2, Q20.3, Q20.4, Q20.5, Q20.6, Q20.8, Q20.9, Q21.0, Q21.1, Q21.2, Q21.3, Q21.4, Q21.8, Q21.9, Q22.0, Q22.1, Q22.2, Q22.3, Q22.4, Q22.5, Q22.6, Q22.8, Q22.9, Q23.0, Q23.1, Q23.2, Q23.3, Q23.4, Q23.8, Q23.9, Q24.0, Q24.1, Q24.2, Q24.3, Q24.4, Q24.5, Q24.6, Q24.8, Q24.9, Q25.0, Q25.1, Q25.2, Q25.3, Q25.4, Q25.5, Q25.6, Q25.7, Q25.8, Q25.9 |
| Atheriosclerosis | I69.8, I70.0, I70.1, I70.2, I70.8, I70.9 |
| Pulmunary Disease | J43.1, J43.2, J43.8, J43.9, J44.00, J44.01, J44.02, J44.03, J44.09, J44.10, J44.11, J44.12, J44.13, J44.19, J44.80, J44.81, J44.82, J44.83, J44.89, J44.90, J44.91, J44.92, J44.93, J44.99, J45.0, J45.1, J45.8, J45.9 |
| Kidney disease | N00,0, N00.1, N00.2, N00.3, N00.4, N00.5, N00.6, N00.7, N00.8, N00.9, N01.0, N01.1, N01.2, N01.3, N01.4, N01.5, N01.6, N01.7, N01.8, N01.9, N02.0, N02.1, N02.2, N02.3, N02.4, N02.5, N02.6, N02.7, N02.8, N02.9, N03.0, N03.1, N03.2, N03.3, N03.4, N03.5, N03.6, N03.7, N03.8, N03.9, N04.0, N04.1, N04.2, N04.3, N04.4, N04.5, N04.6, N04.7, N04.8, N04.9, N05.0, N05.1, N05.2, N05.3, N05.4, N05.5, N05.6, N05.7, N05.8, N05.9, N06.0, N06.1, N06.2, N06.3, N06.4, N06.5, N06.6, N06.7, N06.8, N06.9, N07.0, N07.1, N07.2, N07.3, N07.4, N07.5, N07.6, N07.7, N07.8, N07.9, N08.0, N08.1, N08.2, N08.3, N08.4, N08.5, N08.8, N10, N11.0, N11.1, N11.8, N11.9, N12, N13.0, N13.1, N13.2, N13.3, N13.4, N13.5, N13.6, N13.7, N13.8, N13.9, N14.0, N14.1, N14.2, N14.3, N14.4, N15.0, N15.1, N15.8, N15.9, N16.0, N16.1, N16.2, N16.3, N16.4, N16.5, N16.8, N17.0, N17.1, N17.2, N17.8, N17.9, N18.1, N18.2, N18.3, N18.4, N18.5, N18.9, N19, N20.0 |
| Ischemic CMP | I20.0, I20.1, I20.8, I20.9, I21.0, I21.1, I21.2, I21.3, I21.4, I21.9, I22.0, I22.1, I22.8, I22.9, I23.0, I23.1, I23.2, I23.3, I23.4, I23.5, I23.6, I23.8, I24.0, I24.1, I24.8, I24.9, I25.0, I25.1, I25.2, I25.3, I25.4, I25.5, I25.6, I25.8, I25.9 |
| Malignant diseases | C00.0, C00.1, C00.2, C00.3, C00.4, C00.5, C00.6, C00.8, C00.9, C01, C02.0, C02.1, C02.2, C02.3, C02.4, C02.8, C02.9, C03.0, C03.1, C03.9, C04.0, C04.1, C04.8, C04.9, C05.0, C05.1, C05.2, C05.8, C05.9, C06.0, C06.1, C06.2, C06.8, C06.9, C07 , C08.0, C08.1, C08.8, C08.9, C09.0, C09.1, C09.8, C09.9, C10.0, C10.1, C102., C10.3, C10.4, C10.8, C10.9, C11.0, C11.1, C11.2, C11.3, C11.8, C11.9, C12, C13.0, C13.1, C13.2, C13.8, C13.9, C14.0, C14.2, C14.8, C15.0, C15.1, C15.2, C15.3, C15.4, C15.5, C15.8, C15.9, C16.0, C16.1, C16.2, C16.3, C16.4, C16.5, C16.6, C16.8, C16.9, C17.0, C17.1, C17.2, C17.3, C17.8, C17.9, C18.0, C180.1,  C18.02, C180.3, C18.04, C18.1, C18.11, C18.12, C18.13, C18.14, C182., C18.21, C18.22, C18.23, C18.24, C18.3, C18.31, C18.32, C18.33, C18.34, C18.4, C18.41, C18.42, C18.43, C18.44, C18.5, C18.51, C18.52, C18.53, C18.54, C18.6, C18.61, C18.62, C18.63, C18.64, C18.7, C18.71, C18.72, C18.73, C18.74, C18.8, C18.81, C18.82, C18.83, C18.84, C18.9, C18.91, C18.92, C18.93, C18.94, C19, C19.1, C19.2, C19.3, C19.4, C20, C20.1, C20.2, C20.3, C20.4, C21.0, C21.1, C21.2, C21.8, C22.0, C22.1, C22.2, C22.3, C22.4, C22.7, C22.9, C23, C24.0, C24.1, C24.8, C24.9, C25.0, C25.1, C25.2, C25.3, C25.4, C25.7, C25.8, C25.9, C26.0, C26.1, C26.8, C26.9, C30.0, C30.1, C31.0, C31.1, C31.2, C31.3, C31.8, C31.9, C32.0, C32.1, C32.2, C32.3, C32.8, C32.9, C33, C34.0, C34.1, C34.2, C34.3, C34.8, C34.9, C37, C38.0, C38.1, C38.2, C38.3, C38.4, C38.8, C39.0, C39.8, C39.9, C40.0, C40.1, C40.2, C40.3, C40.8, C40.9, C41.0, C41.1, C41.2, C41.3, C41.4, C41.8, C41.9, C43.0, C43.1, C43.2, C43.3, C43.4, C43.5, C43.6, C43.7, C43.8, C43.9, C44.0, C44.1, C44.2, C44.3, C44.4, C44.5, C44.6, C44.7, C44.8, C44.9, C45.0, C45.1, C45.2, C45.7, C45.9, C46.0, C46.1, C46.2, C46.3, C46.7, C46.8, C46.9, C47.0, C47.1, C47.2, C47.3, C47.4, C47.5, C47.6, C47.8, C47.9, C48.0, C48.1, C48.2, C48.8, C49.0, C49.1, C49.2, C49.3, C49.4, C49.5, C49.6, C49.8, C49.9, C50.0, C50.1, C50.2, C50.3, C50.4, C50.5, C50.6, C50.8, C50.9, C51.0, C51.1, C51.2, C51.8, C51.9, C52, C53.0, C53.1, C53.8, C53.9, C54.0, C54.1, C54.2, C54.3, C54.8, C54.9, C55, C56, C57.0, C57.1, C57.2, C57.3, C57.4, C57.7, C57.8, C57.9, C58, C60.0, C60.1, C60.2, C60.8, C609., C61, C62.0, C62.1, C62.9, C63.0, C63.1, C63.2, C63.7, C63.8, C63.9, C64, C65, C66, C67.0, C67.1, C67.2, C67.3, C67.4, C67.5, C67.6, C67.7, C67.8, C67.9, C68.0, C68.1, C68.8, C68.9, C69.0, C69.1, C69.2, C69.3, C69.4, C69.5, C69.6, C69.8, C69.9, C70.0, C70.1, C70.9, C71.0, C71.1, C71.2, C71.3, C71.4, C71.5, C71.6, C71.7, C71.8, C71.9, C72.0, C72.1, C72.2, C72.3, C72.4, C72.5, C72.8, C72.9, C73, C740., C74.1, C749., C75.0, C75.1, C752., C75.3, C75.4, C75.5, C75.8, C75.9, C76.0, C76.1, C76.2, C76.3, C76.4, C76.5, C76.7, C76.8, C77.0, C77.1, C77.2, C77.3, C77.4, C77.5, C77.8, C77.9, C78.0, C78.1, C78.2, C78.3, C78.4, C78.5, C78.6, C78.7, C78.8, C79.0, C79.1, C79.2, C79.3, C79.4, C79.5, C79.6, C79.7, C79.8, C79.9, C80.0, C80.9, C81.0, C81.1, C81.2, C81.3, C81.4, C81.7, C81.9, C82.0, C82.1, C82.2, C82.3, C82.4, C82.5, C82.6, C82.7, C82.9, C83.0, C83.1, C83.3, C83.5, C83.7, C83.8, C83.9, C840., C84.1, C84.4, C845., C84.6, C84.7, C84.8, C84.9, C85.1, C85.2, C85.7, C85.9, C86.0, C86.1, C86.2, C86.3, C86.4, C86.5, C86.6, C88.0, C88.2, C88.3, C88.4, C887., C88.9, C90.0, C90.1, C90.2, C90.3, C91.0, C91.1, C91.3, C91.4, C91.5, C91.6, C91.7, C91.8, C91.9, C92.0, C92.1, C92.2, C92.3, C92.4, C92.5, C92.6, C92.7, C92.8, C92.9, C93.0, C93.1, C93.3, C93.7, C93.9, C94.0, C94.2, C94.3, C94.4, C94.6, C94.7, C95.0, C95.1, C95.7, C95.9, C96.0, C96.2, C96.4, C96.5, C96.6, C96.7, C96.8, C96.9, C97 |
| Stroke before OP | ICD-Codes as in Table 1 |
| Heart failure before OP | ICD-Codes as in Table 1 |
| Heart attack before OP | ICD-Codes as in Table 1 |

### 3.)  Sensitivity analyses by different age groups (65–75 years and >75 years)

#### Table S6. Hazard ratios (HRs) and corresponding 95% confidence intervals (CIs) from Cox regressions for all-cause mortality in all patients as well as in propensity score–matched subgroups of patients ages 65–75 years, separately for the standard and time-dependent models

| Subgroup: Patients between 65 and 75 years | | | | | | | | |
| --- | --- | --- | --- | --- | --- | --- | --- | --- |
|  | Standard Model | | | | Time dependent Analyses | | | |
|  | All patients of subgroup | | Propensity Score Matched | | All patients of subgroup | | Propensity Score Matched | |
|  | HR (95% CI) | p-value | HR (95% CI) | p-value | HR (95% CI) | p-value | HR (95% CI) | p-value |
| Heart valve (sB-AVR vs. TAVR) : | 2.455  (2.209 - 2.728) | <0.001 | 2.368  (2.019 - 2.779) | <0.001 |  |  |  |  |
| Year 1 |  |  |  |  | 1.889  (1.572 - 2.269) | <0.001 | 1.794  (1.324 - 2.430) | <0.001 |
| Year 2 |  |  |  |  | 3.001  (2.307 - 3.904) | <0.001 | 3.201  (1.965 - 5.214) | <0.001 |
| Year 3 |  |  |  |  | 2.602  (1.971 - 3.435) | <0.001 | 2.093  (1.365 - 3.210) | <0.001 |
| Year 4 |  |  |  |  | 3.082  (2.310 - 4.110) | <0.001 | 2.941  (1.901 - 4.550) | <0.001 |
| Year >4 |  |  |  |  | 2.659  (2.197 - 3.217) | <0.001 | 2.551  (1.955 - 3.327) | <0.001 |
| age (per one year increase) | 1.029  (1.015 - 1.043) | <0.001 | 0.982  (0.954 - 1.010) | 0.20 | 1.029  (1.015 - 1.043) | <0.001 | 0.982  (0.954 - 1.010) | 0.20 |
| sex (male vs. female) | 0.871  (0.800 - 0.948) | 0.001 | 0.889  (0.757 - 1.043) | 0.15 | 0.869  (0.798 - 0.946) | 0.001 | 0.889  (0.758 - 1.043) | 0.15 |
| heart failure* | 1.529  (1.378 - 1.698) | <0.001 | 1.574  (1.312 - 1.888) | <0.001 | 1.536  (1.384 - 1.705) | <0.001 | 1.579  (1.316 - 1.895) | <0.001 |
| heart attack* | 1.183  (0.960 - 1.458) | 0.12 | 0.825  (0.466 - 1.463) | 0.51 | 1.173  (0.952 - 1.446) | 0.13 | 0.819  (0.462 - 1.452) | 0.50 |
| stroke* | 1.213  (0.924 - 1.591) | 0.16 | 1.122  (0.550 - 2.292) | 0.75 | 1.205  (0.918 - 1.581) | 0.18 | 1.131  (0.553 - 2.310) | 0.74 |
| diabetes mellitus* | 1.461  (1.326 - 1.609) | <0.001 | 1.337  (1.118 - 1.600) | 0.001 | 1.464  (1.329 - 1.613) | <0.001 | 1.342  (1.122 - 1.606) | 0.001 |
| adiposity* | 1.033  (0.899 - 1.186) | 0.65 | 1.010  (0.767 - 1.329) | 0.95 | 1.030  (0.897 - 1.182) | 0.68 | 1.008  (0.766 - 1.327) | 0.95 |
| hyperlipidemia* | 0.834  (0.758 - 0.918) | <0.001 | 0.723  (0.593 - 0.880) | 0.001 | 0.832  (0.756 - 0.916) | <0.001 | 0.719  (0.590 - 0.877) | 0.001 |
| hyperuricemia/gout* | 1.159  (0.957 - 1.404) | 0.13 | 1.273  (0.832 - 1.947) | 0.27 | 1.160  (0.957 - 1.405) | 0.13 | 1.281  (0.837 - 1.960) | 0.25 |
| valvular, rhythmological and other CMPs* | 0.906  (0.797 - 1.031) | 0.14 | 1.015  (0.757 - 1.362) | 0.92 | 0.907  (0.797 - 1.032) | 0.14 | 1.017  (0.758 - 1.364) | 0.91 |
| iCMP* | 1.093  (1.003 - 1.192) | 0.04 | 1.135  (0.963 - 1.337) | 0.13 | 1.094  (1.003 - 1.193) | 0.04 | 1.134  (0.963 - 1.337) | 0.13 |
| atherosclerosis* | 1.220  (1.046 - 1.423) | 0.01 | 1.149  (0.827 - 1.596) | 0.41 | 1.218  (1.044 - 1.421) | 0.01 | 1.150  (0.828 - 1.597) | 0.41 |
| pulmonary diseases* | 1.141  0.934 - 1.394) | 0.20 | 1.119  (0.745 - 1.681) | 0.59 | 1.155  (0.945 - 1.411) | 0.16 | 1.126  (0.749 - 1.692) | 0.57 |
| kidney diseases* | 1.565  (1.391 - 1.760) | <0.001 | 1.583  (1.296 - 1.934) | <0.001 | 1.564  (1.391 - 1.759) | <0.001 | 1.584  (1.296 - 1.935) | <0.001 |
| malignant diseases* | 1.592  (1.372 - 1.847) | <0.001 | 1.709  (1.283 - 2.276) | <0.001 | 1.598  (1.377 - 1.853) | <0.001 | 1.717  (1.289 - 2.288) | <0.001 |

#### Table S7. Baseline characteristics between groups before and after propensity score matching for the subgroup of patients ages 65 to 75 years

| **Subgroup: Patients between 65 and 75 years** | | | | | | |
| --- | --- | --- | --- | --- | --- | --- |
|  | **All Data** | | | **After PSM** | | |
|  | **Bio**  **(n = 6483)** | **TAVR**  **(n = 1092)** | **p-value** | **Bio**  **(n = 869)** | **TAVR**  **(n = 869)** | **p-value** |
| **Age**  **(median,** | 71 (68 - 73) | 73 (70 - 74) | <0.001 | 73 (70 - 74) | 73 (70 - 74) | 0.77 |
| **Sex** | | | | | | |
| **M** | 4090 (63.09%) | 638 (58.42%) |  | 519 (59.72%) | 521 (59.95%) |  |
| F | 2393 (36.91%) | 454 (41.58%) | 0.004 | 350 (40.28%) | 348 (40.05%) | 0.96 |
| **Heart failure** | | | | | | |
| **No** | 5703 (87.97%) | 809 (74.08%) |  | 712 (81.93%) | 699 (80.44%) |  |
| **Yes** | 780 (12.03%) | 283 (25.92%) | <0.001 | 157 (18.07%) | 170 (19.56%) | 0.46 |
| **Heart attack** | | | | | | |
| **No** | 6274 (96.78%) | 1064 (97.44%) |  | 855 (98.39%) | 851 (97.93%) |  |
| **Yes** | 209 (3.22%) | 28 (2.56%) | 0.29 | 14 (1.61%) | 18 (2.07%) | 0.59 |
| **Stroke** | | | | | | |
| **No** | 6368 (98.23%) | 1069 (97.89%) |  | 861 (99.08%) | 858 (98.73%) |  |
| **Yes** | 115 (1.77%) | 23 (2.11%) | 0.52 | 8 (0.92%) | 11 (1.27%) | 0.65 |
| **Diabetes mellitus** | | | | | | |
| **No** | 5229 (80.66%) | 759 (69.51%) |  | 663 (76.29%) | 652 (75.03%) |  |
| **Yes** | 1254 (19.34%) | 333 (30.49%) | <0.001 | 206 (23.71%) | 217 (24.97%) | 0.58 |
| **Adiposity** | | | | | | |
| **No** | 5953 (91.82%) | 965 (88.37%) |  | 805 (92.64%) | 789 (90.79%) |  |
| **Yes** | 530 (8.18%) | 127 (11.63%) | <0.001 | 64 (7.36%) | 80 (9.21%) | 0.19 |
| **Hyperlipidemia** | | | | | | |
| **No** | 4685 (72.27%) | 798 (73.08%) |  | 670 (77.1%) | 662 (76.18%) |  |
| **Yes** | 1798 (27.73%) | 294 (26.92%) | 0.60 | 199 (22.9%) | 207 (23.82%) | 0.69 |
| **Hyperuricemia/gout** | | | | | | |
| **No** | 6255 (96.48%) | 1039 (95.15%) |  | 848 (97.58%) | 844 (97.12%) |  |
| **Yes** | 228 (3.52%) | 53 (4.85%) | 0.04 | 21 (2.42%) | 25 (2.88%) | 0.65 |
| **Valvular, rhythmological and other CMPs** | | | | | | |
| **No** | 958 (14.78%) | 82 (7.51%) |  | 71 (8.17%) | 77 (8.86%) |  |
| **Yes** | 5525 (85.22%) | 1010 (92.49%) | <0.001 | 798 (91.83%) | 792 (91.14%) | 0.67 |
| **Ischemic CMP** | | | | | | |
| **No** | 3041 (46.91%) | 464 (42.49%) |  | 388 (44.65%) | 388 (44.65%) |  |
| **Yes** | 3442 (53.09%) | 628 (57.51%) | 0.007 | 481 (55.35%) | 481 (55.35%) | 1.00 |
| **Atherosclerosis** | | | | | | |
| **No** | 6142 (94.74%) | 995 (91.12%) |  | 833 (95.86%) | 822 (94.59%) |  |
| **Yes** | 341 (5.26%) | 97 (8.88%) | <0.001 | 36 (4.14%) | 47 (5.41%) | 0.26 |
| **Pulmonary diseases** | | | | | | |
| **No** | 6259 (96.54%) | 984 (90.11%) |  | 826 (95.05%) | 832 (95.74%) |  |
| **Yes** | 224 (3.46%) | 108 (9.89%) | <0.001 | 43 (4.95%) | 37 (4.26%) | 0.57 |
| **Kidney diseases** | | | | | | |
| **0: No** | 5906 (91.1%) | 851 (77.93%) |  | 747 (85.96%) | 745 (85.73%) |  |
| **Yes** | 577 (8.9%) | 241 (22.07%) | <0.001 | 122 (14.04%) | 124 (14.27%) | 0.95 |
| **Malignant diseases** | | | | | | |
| **No** | 6180 (95.33%) | 1007 (92.22%) |  | 821 (94.48%) | 822 (94.59%) |  |
| **Yes** | 303 (4.67%) | 85 (7.78%) | <0.001 | 48 (5.52%) | 47 (5.41%) | 1.00 |

#### Table S8. Hazard ratios (HRs) and corresponding 95% confidence intervals (CIs) from Cox regressions for all-cause mortality in all patients as well as in propensity score–matched subgroups of patients older than 75 years, separately for the standard and time-dependent models

| Subgroup: Patients older than 75 years | | | | | | | | |
| --- | --- | --- | --- | --- | --- | --- | --- | --- |
|  | Standard Model | | | | Time dependent Analyses | | | |
|  | All patients of subgroup | | Propensity Score Matched | | All patients of subgroup | | Propensity Score Matched | |
|  | HR (95% CI) | p-value | HR (95% CI) | p-value | HR (95% CI) | p-value | HR (95% CI) | p-value |
| Heart valve (sB-AVR vs. TAVR) : | 1.354  (1.272 - 1.442) | <0.001 | 1.368  (1.259 - 1.485) | <0.001 |  |  |  |  |
| Year 1 |  |  |  |  | 0.919  (0.821 - 1.028) | 0.14 | 0.888  (0.755 - 1.044) | 0.15 |
| Year 2 |  |  |  |  | 1.699  (1.439 - 2.006) | <0.001 | 1.852  (1.446 - 2.372) | <0.001 |
| Year 3 |  |  |  |  | 1.655  (1.394 - 1.966) | <0.001 | 1.793  (1.404 - 2.29) | <0.001 |
| Year 4 |  |  |  |  | 1.542  (1.306 - 1.821) | <0.001 | 1.538  (1.225 - 1.931) | <0.001 |
| Year >4 |  |  |  |  | 1.473  (1.342 - 1.616) | <0.001 | 1.478  (1.295 - 1.686) | <0.001 |
| age (per one year increase) | 1.064  (1.056 - 1.072) | <0.001 | 1.054  (1.041 - 1.068) | <0.001 | 1.065  (1.057 - 1.073) | <0.001 | 1.055  (1.041 - 1.068) | <0.001 |
| sex (male vs. female) | 0.863  (0.817 - 0.912) | <0.001 | 0.889  (0.820 - 0.964) | 0.004 | 0.862  (0.816 - 0.911) | <0.001 | 0.889  (0.820 - 0.963) | 0.004 |
| heart failure* | 1.377  (1.289 - 1.472) | <0.001 | 1.338  (1.197 - 1.496) | <0.001 | 1.380  (1.291 - 1.475) | <0.001 | 1.339  (1.198 - 1.497) | <0.001 |
| heart attack* | 1.149  (0.988 - 1.336) | 0.07 | 1.364  (0.962 - 1.933) | 0.08 | 1.144  (0.984 - 1.330) | 0.08 | 1.365  (0.963 - 1.935) | 0.08 |
| stroke* | 1.175  (0.958 - 1.440) | 0.12 | 1.391  (0.802 - 2.410) | 0.24 | 1.172  (0.956 - 1.437 | 0.13 | 1.389  (0.801 - 2.408) | 0.24 |
| diabetes mellitus* | 1.330  1.239 - 1.427) | <0.001 | 1.391  (1.238 - 1.563) | <0.001 | 1.331  (1.240 - 1.428) | <0.001 | 1.390  (1.237 - 1.562) | <0.001 |
| adiposity* | 1.128  (0.998 - 1.276) | 0.05 | 1.036  (0.787 - 1.364) | 0.8 | 1.127  (0.996 - 1.274) | 0.06 | 1.038  (0.788 - 1.367) | 0.79 |
| hyperlipidemia* | 0.822  (0.769 - 0.879) | <0.001 | 0.808  (0.729 - 0.895) | <0.001 | 0.823  (0.770 - 0.880) | <0.001 | 0.808  (0.729 - 0.895) | <0.001 |
| hyperuricemia/gout* | 1.067  (0.927 - 1.228) | 0.37 | 1.340  (0.978 - 1.836) | 0.07 | 1.068  (0.928 - 1.229) | 0.36 | 1.341  (0.979 - 1.837) | 0.07 |
| valvular, rhythmological and other CMPs* | 0.840  (0.768 - 0.919) | <0.001 | 0.821  (0.726 - 0.929) | 0.002 | 0.841  (0.769 - 0.920) | <0.001 | 0.822  (0.726 - 0.929) | 0.002 |
| iCMP* | 1.069  (1.010 - 1.133) | 0.02 | 1.105  (1.016 - 1.202) | 0.02 | 1.070  (1.010 - 1.133) | 0.02 | 1.105  (1.016 - 1.202) | 0.02 |
| atherosclerosis* | 1.183  (1.073 - 1.305) | <0.001 | 1.261  (1.035 - 1.537) | 0.02 | 1.183  (1.072 - 1.304) | <0.001 | 1.259  (1.033 - 1.534) | 0.02 |
| pulmonary diseases* | 1.364  (1.184 - 1.571) | <0.001 | 1.294  (0.945 - 1.772) | 0.11 | 1.384  (1.201 - 1.595) | <0.001 | 1.300  (0.949 - 1.780) | 0.10 |
| kidney diseases* | 1.344  (1.249 - 1.447) | <0.001 | 1.448  (1.275 - 1.644) | <0.001 | 1.347  (1.252 - 1.449) | <0.001 | 1.448  (1.275 - 1.645) | <0.001 |
| malignant diseases* | 1.355  (1.217 - 1.507) | <0.001 | 1.307  (1.063 - 1.607) | 0.01 | 1.358  (1.221 - 1.512) | <0.001 | 1.309  (1.064 - 1.609) | 0.01 |

#### Table S9. Baseline characteristics between groups before and after propensity score matching for the subgroup of patients older than 75 years

| **Subgroup: Patients older than 75 years** | | | | | | |
| --- | --- | --- | --- | --- | --- | --- |
|  | **All Data** | | | **After PSM** | | |
|  | **Bio**  **(n = 5266)** | **TAVI**  **(n = 6041)** | **p-value** | **Bio**  **(n = 2764)** | **TAVR**  **(n = 2764)** | **p-value** |
| **Age**  **(median,** | 79 (77 - 81) | 83 (80 - 86) | <0.001 | 80 (78 - 83) | 80 (78 - 83) | 0.88 |
| **Sex** | | | | | | |
| **M** | 2796 (53.1%) | 2539 (42.03%) |  | 1252 (45.3%) | 1243 (44.97%) |  |
| F | 2470 (46.9%) | 3502 (57.97%) | <0.001 | 1512 (54.7%) | 1521 (55.03%) | 0.83 |
| **Heart failure** | | | | | | |
| **No** | 4512 (85.68%) | 4628 (76.61%) |  | 2401 (86.87%) | 2413 (87.3%) |  |
| **Yes** | 754 (14.32%) | 1413 (23.39%) | <0.001 | 363 (13.13%) | 351 (12.7%) | 0.66 |
| **Heart attack** | | | | | | |
| **No** | 5058 (96.05%) | 5925 (98.08%) |  | 2737 (99.02%) | 2736 (98.99%) |  |
| **Yes** | 208 (3.95%) | 116 (1.92%) | <0.001 | 27 (0.98%) | 28 (1.01%) | 1.00 |
| **Stroke** | | | | | | |
| **No** | 5189 (98.54%) | 5928 (98.13%) |  | 2755 (99.67%) | 2750 (99.49%) |  |
| **Yes** | 77 (1.46%) | 113 (1.87%) | 0.11 | 9 (0.33%) | 14 (0.51%) | 0.40 |
| **Diabetes mellitus** | | | | | | |
| **No** | 4403 (83.61%) | 4959 (82.09%) |  | 2420 (87.55%) | 2423 (87.66%) |  |
| **Yes** | 863 (16.39%) | 1082 (17.91%) | 0.03 | 344 (12.45%) | 341 (12.34%) | 0.94 |
| **Adiposity** | | | | | | |
| **No** | 5012 (95.18%) | 5753 (95.23%) |  | 2709 (98.01%) | 2706 (97.9%) |  |
| **Yes** | 254 (4.82%) | 288 (4.77%) | 0.92 | 55 (1.99%) | 58 (2.1%) | 0.5 |
| **Hyperlipidemia** | | | | | | |
| **No** | 3864 (73.38%) | 4568 (75.62%) |  | 2157 (78.04%) | 2135 (77.24%) |  |
| **Yes** | 1402 (26.62%) | 1473 (24.38%) | 0.007 | 607 (21.96%) | 629 (22.76%) | 0.50 |
| **Hyperuricemia/gout** | | | | | | |
| **No** | 5070 (96.28%) | 5812 (96.21%) |  | 2729 (98.73%) | 2717 (98.3%) |  |
| **Yes** | 196 (3.72%) | 229 (3.79%) | 0.89 | 35 (1.27%) | 47 (1.7%) | 0.22 |
| **Valvular, rhythmological and other CMPs** | | | | | | |
| **No** | 736 (13.98%) | 524 (8.67%) |  | 342 (12.37%) | 339 (12.26%) |  |
| **Yes** | 4530 (86.02%) | 5517 (91.33%) | <0.001 | 2422 (87.63%) | 2425 (87.74%) | 0.94 |
| **Ischemic CMP** | | | | | | |
| **No** | 2344 (44.51%) | 2915 (48.25%) |  | 1336 (48.34%) | 1337 (48.37%) |  |
| **Yes** | 2922 (55.49%) | 3126 (51.75%) | <0.001 | 1428 (51.66%) | 1427 (51.63%) | 1.00 |
| **Atherosclerosis** | | | | | | |
| **No** | 4913 (93.3%) | 5536 (91.64%) |  | 2657 (96.13%) | 2656 (96.09%) |  |
| **Yes** | 353 (6.7%) | 505 (8.36%) | 0.001 | 107 (3.87%) | 108 (3.91%) | 1.00 |
| **Pulmonary diseases** | | | | | | |
| **No** | 5139 (97.59%) | 5694 (94.26%) |  | 2723 (98.52%) | 2712 (98.12%) |  |
| **Yes** | 127 (2.41%) | 347 (5.74%) | <0.001 | 41 (1.48%) | 52 (1.88%) | 0.30 |
| **Kidney diseases** | | | | | | |
| **0: No** | 4640 (88.11%) | 4804 (79.52%) |  | 2491 (90.12%) | 2489 (90.05%) |  |
| **Yes** | 626 (11.89%) | 1237 (20.48%) | <0.001 | 273 (9.88%) | 275 (9.95%) | 0.96 |
| **Malignant diseases** | | | | | | |
| **No** | 5014 (95.21%) | 5675 (93.94%) |  | 2682 (97.03%) | 2678 (96.89%) |  |
| **Yes** | 252 (4.79%) | 366 (6.06%) | 0.003 | 82 (2.97%) | 86 (3.11%) | 0.81 |

### 4.) Sensitivity analyses: by different years of the index procedure

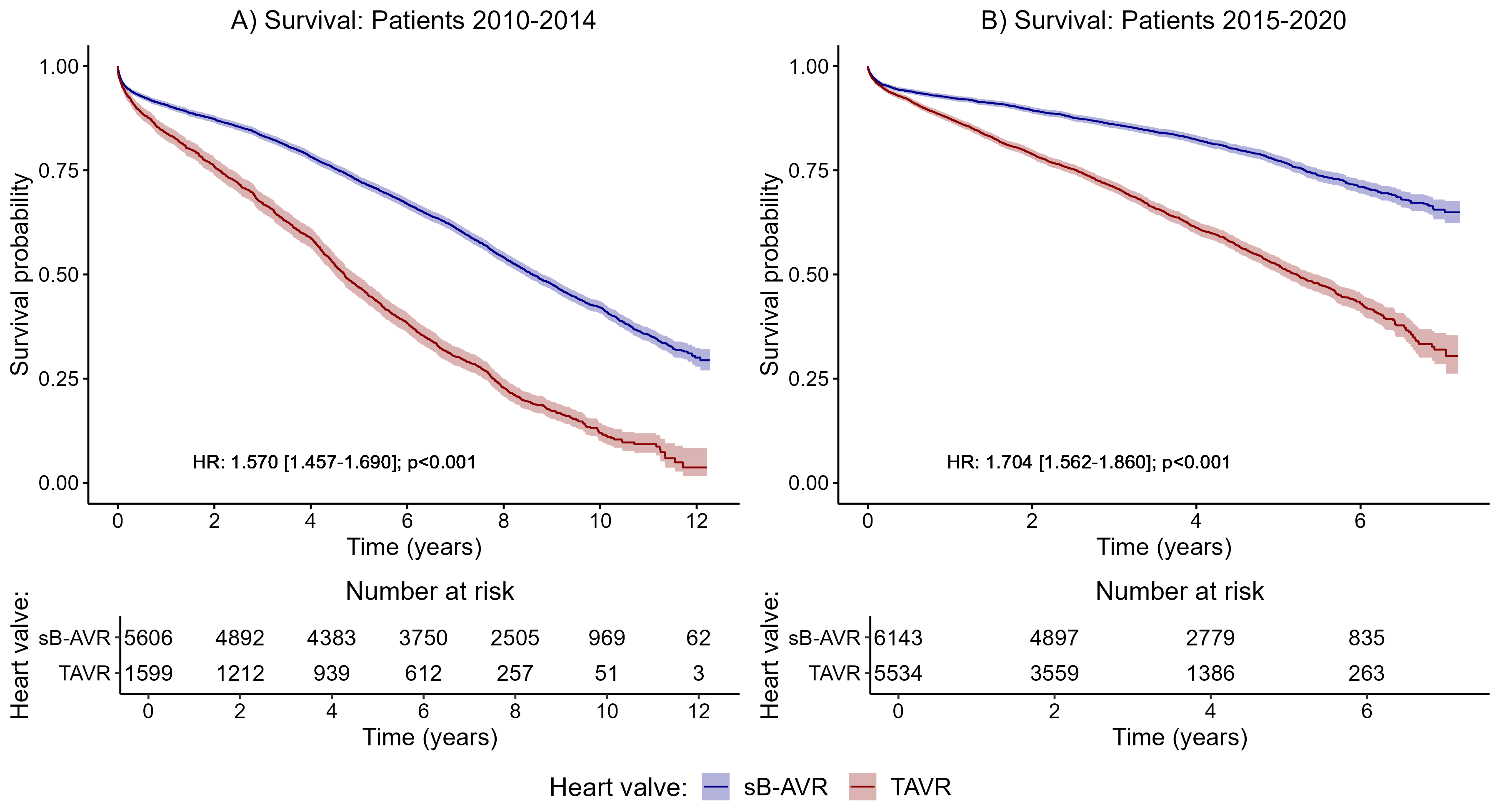

Supplementary Figure 1. Survival probabilities and corresponding 95% confidence intervals, separately for the two investigated groups (red: TAVR; blue: sB-AVR) within different subgroups of patients: A) with index procedure before 2015 and B) with index procedure during or after 2015.

#### Table S10. Hazard ratios (HRs) with corresponding 95% confidence intervals from Cox regressions for all-cause mortality in the subgroup of patients with index-op before 2015 and during/after 2015

|  | Patients 2010 - 2014 | | Patients 2015 - 2020 | |
| --- | --- | --- | --- | --- |
|  | HR (95% CI) | p-value | HR (95% CI) | p-value |
| Heart valve (sB-AVR vs. TAVR) | 1.570 (1.457 - 1.690) | <0.001 | 1.704 (1.562 - 1.860) | <0.001 |
| age (per one year increase) | 1.059 (1.053 - 1.065) | <0.001 | 1.042 (1.035 - 1.049) | <0.001 |
| sex (male vs. female) | 0.891 (0.839 - 0.947) | <0.001 | 0.808 (0.752 - 0.869) | <0.001 |
| heart failure before OP | 1.344 (1.247 - 1.449) | <0.001 | 1.487 (1.366 - 1.619) | <0.001 |
| heart attack before OP | 1.114 (0.955 - 1.301) | 0.17 | 1.227 (1.005 - 1.498) | 0.04 |
| stroke before OP | 1.063 (0.846 - 1.335) | 0.60 | 1.409 (1.116 - 1.779) | 0.004 |
| diabetes mellitus* | 1.387 (1.288 - 1.494) | <0.001 | 1.351 (1.237 - 1.476) | <0.001 |
| adiposity* | 1.133 (1.007 - 1.273) | 0.04 | 1.082 (0.935 - 1.252) | 0.29 |
| hyperlipidemia* | 0.851 (0.793 - 0.913) | <0.001 | 0.782 (0.717 - 0.853) | <0.001 |
| hyperuricemia/gout* | 1.181 (1.014 - 1.376) | 0.03 | 1.001 (0.846 - 1.185) | 1.00 |
| valvular, rhythmological and other CMPs* | 0.910 (0.827 - 1.001) | 0.05 | 0.793 (0.706 - 0.891) | <0.001 |
| iCMP* | 1.117 (1.049 - 1.190) | <0.001 | 1.022 (0.950 - 1.099) | 0.57 |
| atherosclerosis* | 1.182 (1.050 - 1.330) | 0.01 | 1.222 (1.089 - 1.372) | <0.001 |
| pulmonary diseases* | 1.295 (1.024 - 1.639) | 0.03 | 1.371 (1.199 - 1.569) | <0.001 |
| kidney diseases* | 1.376 (1.259 - 1.504) | <0.001 | 1.481 (1.357 - 1.618) | <0.001 |
| malignant diseases* | 1.330 (1.180 - 1.498) | <0.001 | 1.534 (1.352 - 1.741) | <0.001 |

### 5.)  Sensitivity analyses: by comorbid diseases

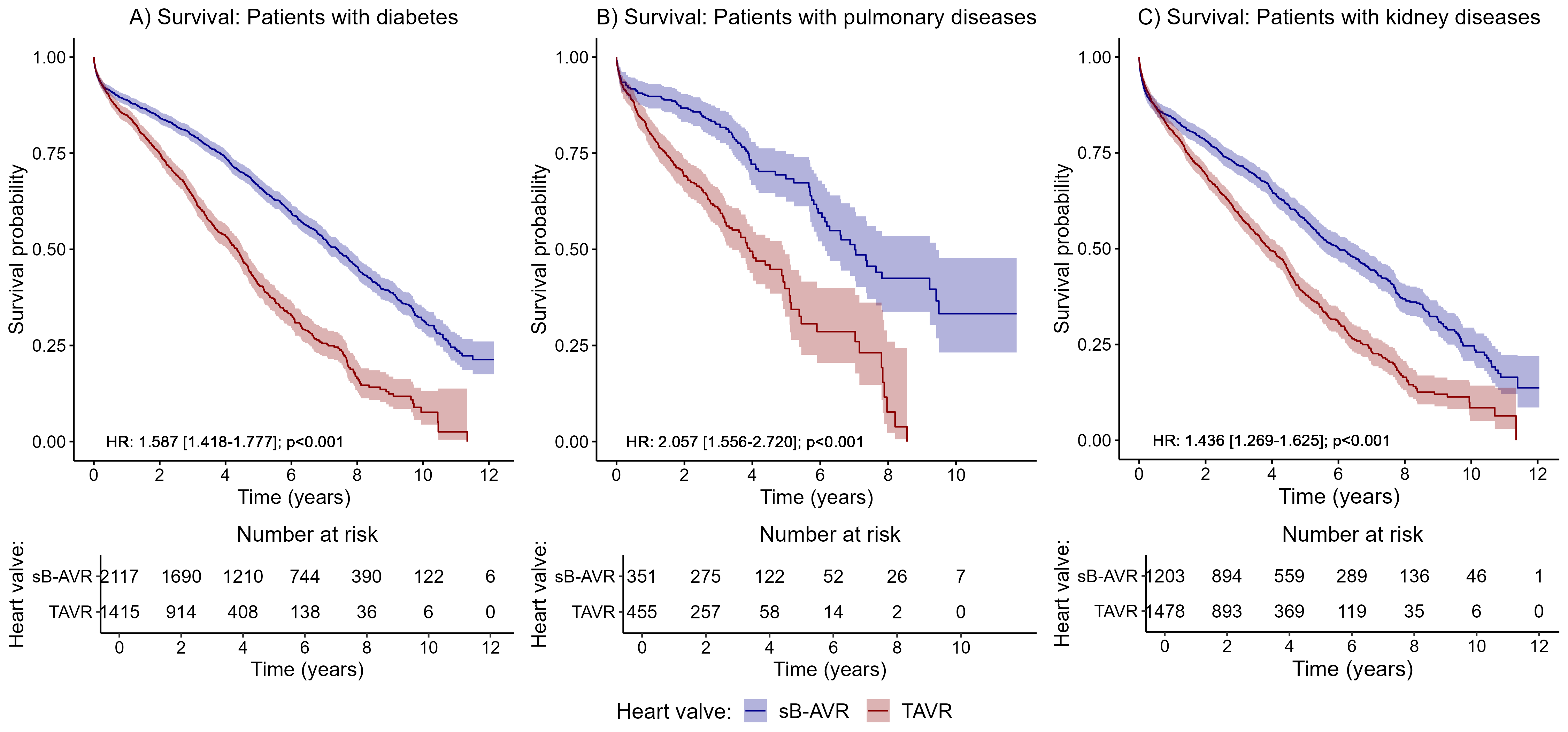

Supplementary Figure 2. Survival probabilities and corresponding 95% confidence intervals, separately for the two investigated groups (red: TAVR; blue: sB-AVR) within different subgroups of patients: A) with diabetes, B) with pulmonary diseases, and C) with kidney diseases.

#### Table S11. Hazard ratios (HRs) and corresponding 95% confidence intervals (CIs) from Cox regressions for all-cause mortality in subgroups of patients with diabetes, pulmonary diseases, and kidney diseases

|  | Patients with diabetes | | Patients with pulmonary diseases | | Patients with kidney diseases | |
| --- | --- | --- | --- | --- | --- | --- |
|  | HR (95% CI) | p-value | HR (95% CI) | p-value | HR (95% CI) | p-value |
| Heart valve (sB-AVR vs. TAVR) | 1.587 (1.418 - 1.777) | <0.001 | 2.057 (1.556 - 2.72) | <0.001 | 1.436 (1.269 - 1.625) | <0.001 |
| age (per one year increase) | 1.038 (1.028 - 1.048) | <0.001 | 1.029 (1.008 - 1.051 | 0.008 | 1.025 (1.015 - 1.036) | <0.001 |
| sex (male vs. female) | 0.849 (0.769 - 0.938) | 0.001 | 0.812 (0.643 - 1.026) | 0.08 | 0.853 (0.765 - 0.952) | 0.004 |
| heart failure before OP | 1.565 (1.412 - 1.735) | <0.001 | 1.381 (1.086 - 1.756) | 0.008 | 1.43 (1.285 - 1.592) | <0.001 |
| heart attack before OP | 1.114 (0.901 - 1.377) | 0.32 | 1.186 (0.672 - 2.094) | 0.56 | 0.989 (0.769 - 1.272) | 0.93 |
| stroke before OP | 1.333 (1.01 - 1.76) | 0.04 | 0.869 (0.376 - 2.008) | 0.74 | 1.144 (0.823 - 1.591) | 0.42 |
| diabetes mellitus* |  |  | 1.32 (1.019 - 1.711) | 0.04 | 1.243 (1.109 - 1.393) | <0.001 |
| adiposity* | 1.089 (0.956 - 1.24) | 0.20 | 0.952 (0.656 - 1.381) | 0.79 | 1.036 (0.878 - 1.223) | 0.68 |
| hyperlipidemia* | 0.926 (0.839 - 1.022) | 0.13 | 0.88 (0.695 - 1.114) | 0.29 | 0.829 (0.739 - 0.93) | 0.001 |
| hyperuricemia/gout* | 1.157 (0.962 - 1.391) | 0.12 | 1.027 (0.686 - 1.537) | 0.90xwxw | 1.127 (0.957 - 1.328) | 0.15 |
| valvular, rhythmological and other CMPs* | 0.731 (0.431 - 1.24) | 0.24 | 1.122 (0.277 - 4.552) | 0.87 | 0.916 (0.548 - 1.531) | 0.74 |
| iCMP* | 1.088 (0.98 - 1.208) | 0.12 | 0.949 (0.743 - 1.212) | 0.67 | 1.107 (0.987 - 1.241) | 0.08 |
| atherosclerosis* | 1.217 (1.05 - 1.409) | 0.009 | 1.441 (1.06 - 1.959) | 0.02 | 1.193 (1.026 - 1.388) | 0.02 |
| pulmonary diseases* | 1.23 (0.995 - 1.52) | 0.06 |  |  | 1.108 (0.903 - 1.359) | 0.33 |
| kidney diseases* | 1.349 (1.208 - 1.507) | <0.001 | 1.236 (0.954 - 1.6) | 0.11 |  |  |
| malignant diseases* | 1.236 (1.042 - 1.467) | 0.02 | 1.348 (0.935 - 1.945) | 0.11 | 1.268 (1.071 - 1.502) | 0.006 |

### 6.)  Sensitivity analyses: by patients with and without a pacemaker implantation at 1 month after index procedure

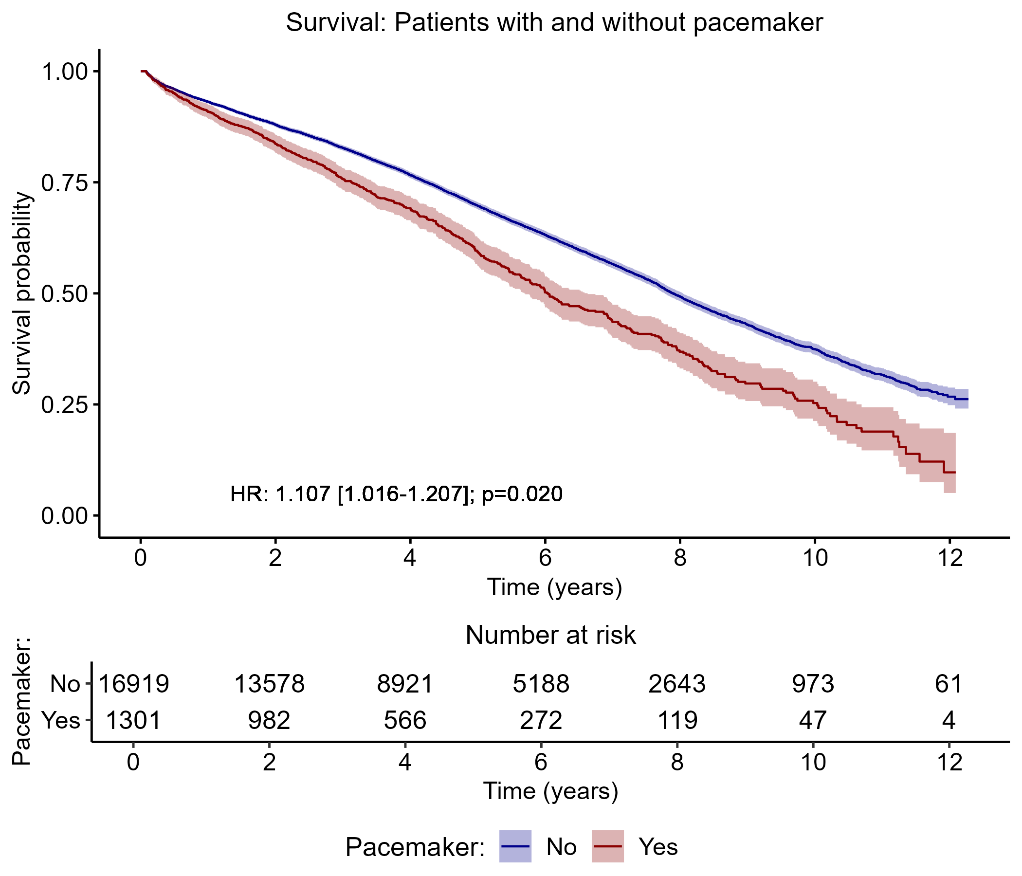

#### Supplementary Figure 3. Survival probabilities and corresponding 95% confidence intervals, separately for patients receiving and not receiving a pacemaker 1 month after index procedure.

#### Table S12. Hazard ratios (HRs) with corresponding 95% confidence intervals from Cox regressions for all-cause mortality for the sensitivity analysis for patients receiving and not receiving a pacemaker 1 month after the index procedure

|  | All-Cause Mortality | |
| --- | --- | --- |
|  | HR (95% CI) | p-value |
| Heart valve (sB-AVR vs. TAVI) | 1.667 (1.573 – 1.767) | <0.001 |
| age (per one year increase) | 1.053 (1.048 – 1.058) | <0.001 |
| sex (male vs. female) | 0.845 (0.805 – 0.887) | <0.001 |
| heart failure before OP | 1.408 (1.328 – 1.493) | <0.001 |
| heart attack before OP | 1.146 (1.007 – 1.303) | 0.04 |
| Stroke before OP | 1.195 (1.006 – 1.418) | 0.04 |
| diabetes mellitus | 1.393 (1.313 – 1.478) | <0.001 |
| Adiposity | 1.103 (1.002 – 1.214) | 0.04 |
| Hyperlipidemia | 0.825 (0.779 – 0.873) | <0.001 |
| hyperuricemia/gout | 1.038 (0.920 – 1.172) | 0.55 |
| valvular, rhythmological and other CMPs | 0.909 (0.840 – 0.982) | 0.02 |
| iCMP | 1.092 (1.038 – 1.147) | <0.001 |
| Atherosclerosis | 1.208 (1.108 – 1.316) | <0.001 |
| pulmonary diseases | 1.333 (1.180 – 1.507) | <0.001 |
| kidney diseases | 1.416 (1.327 – 1.512) | <0.001 |
| malignant diseases | 1.470 (1.344 – 1.607) | <0.001 |
| Pacemaker 1 month after OP | 1.107 (1.016 – 1.207) | 0.02 |

### 7.)  Secondary outcomes

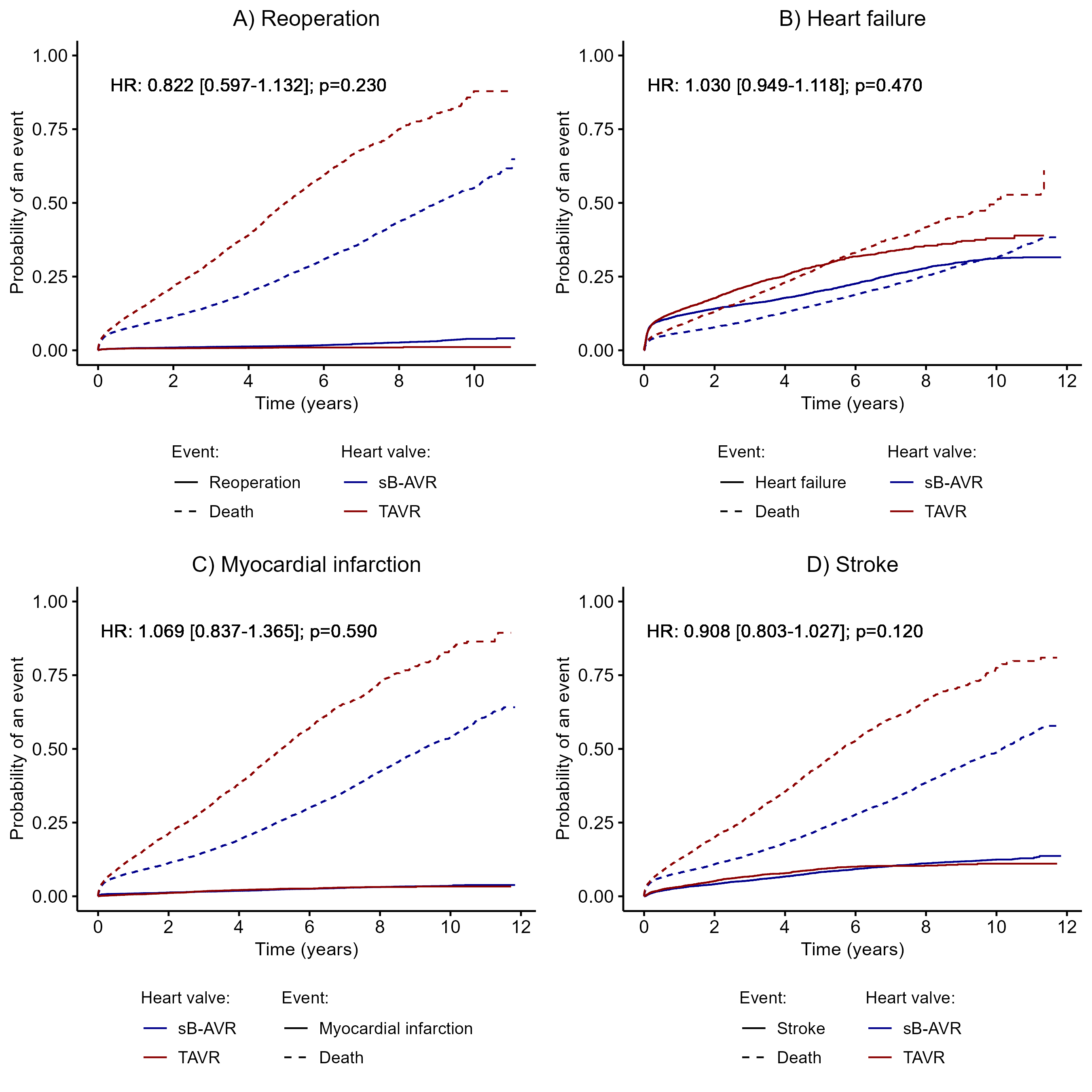

#### Supplementary Figure 4. Cumulative incidence curves for (A) reoperation, (B) heart failure, (C) myocardial infarcion, and (D) stroke, with death as the competing event.

#### Table S13. Hazard ratios (HRs) with corresponding 95% confidence intervals (CIs) for the secondary outcome parameters of reoperation, heart failure, myocardial infarction, and stroke (competing risk analyses)

|  | reoperation | | heart failure | | Myocardial infarction | | stroke | |
| --- | --- | --- | --- | --- | --- | --- | --- | --- |
|  | HR (95% CI) | p-value | HR (95% CI) | p-value | HR (95% CI) | p-value | HR (95% CI) | p-value |
| Heart valve (sB-AVR vs. TAVR) | 0.822  (0.597 – 1.132 | 0.23 | 1.030  (0.949 – 1.118) | 0.47 | 1.069  (0.837 – 1.365) | 0.59 | 0.908  (0.803 – 1.027) | 0.12 |
| age (per one year increase) | 0.920  (0.900 – 0.940 | < 0.001 | 1.030  (1.024 – 1.036) | < 0.001 | 0.989  (0.971 – 1.006) | 0.21 | 1.015  (1.006 – 1.024) | 0.001 |
| sex (male vs. female) | 1.317  (1.036 – 1.675) | 0.03 | 1.082  (1.012 – 1.157) | 0.02 | 1.079  (0.890 – 1.307) | 0.44 | 1.024  (0.926 – 1.131) | 0.65 |
| heart failure* | 0.875  (0.608 – 1.262) | 0.48 | - | - | 0.925  (0.703 – 1.218) | 0.58 | 1.005  (0.877 – 1.151) | 0.95 |
| heart attack* | 0.980  (0.457 – 2.099) | 0.96 | 1.369  (1.140 – 1.644) | 0.001 | 4.282  (3.165.- 5.795) | < 0.001 | 0.889  (0.658 – 1.201) | 0.44 |
| stroke* | 1.381  (0.650 – 2.933) | 0.40 | 1.126  (0.882 – 1.437) | 0.34 | 0.811  (0.365 – 1.801 | 0.61 | 2.490  (1.922 – 3.225) | < 0.001 |
| diabetes mellitus* | 0.719  (0.511 – 1.012) | 0.06 | 1.324  (1.215 – 1.443) | < 0.001 | 1.177  (0.928 – 1.493) | 0.18 | 1.043  (0.916 – 1.188) | 0.52 |
| adiposity* | 1.204  (0.776 – 1.869) | 0.41 | 1.136  (0.992 – 1.301) | 0.07 | 0.930  (0.636 – 1.360) | 0.71 | 0.960  (0.778 – 1.184) | 0.70 |
| hyperlipidemia* | 1.262  (0.953 – 1.671) | 0.10 | 0.818  (0.754 – 0.886) | < 0.001 | 0.932  (0.745 – 1.166) | 0.54 | 1.129  (1.008 – 1.264) | 0.04 |
| hyperuricemia/gout* | 1.159  (0.624 – 2.153) | 0.64 | 1.183  (0.990 – 1.413) | 0.06 | 0.917  (0.545 – 1.542) | 0.740 | 0.885  (0.672 – 1.165) | 0.38 |
| valvular, rhythmological and other CMPs* | 0.823  (0.592 – 1.144) | 0.25 | 0.914  (0.829 – 1.008) | 0.07 | 0.553  (0.426 – 0.718) | < 0.001 | 1.094  (0.926 – 1.291) | 0.29 |
| iCMP* | 0.651  (0.508 – 0.835) | 0.001 | 1.048  (0.978 – 1.122) | 0.18 | 1.575  (1.272 – 1.948) | < 0.001 | 1.058  (0.953 – 1.173) | 0.29 |
| atherosclerosis* | 1.246  (0.770 – 2.019) | 0.37 | 1.269  (1.117 – 1.441) | < 0.001 | 1.436  (1.044 – 1.975) | 0.03 | 1.051  (0.866 – 1.274 | 0.62 |
| pulmonary diseases* | 0.692  (0.305 – 1.573) | 0.38 | 1.176  (0.987 – 1.402) | 0.07 | 0.978  (0.577 – 1.659) | 0.93 | 0.732  (0.537 – 0.997) | 0.05 |
| kidney diseases* | 1.195  (0.817 – 1.748) | 0.36 | 1.329  (1.202 – 1.470) | < 0.001 | 0.978  (727 -– 1.316) | 0.88 | 1.005  (0.864 – 1.170) | 0.95 |
| malignant diseases* | 1.328  (0.813 – 2.171) | 0.26 | 1.010  (0.872 – 1.168 | 0.90 | 0.408  (0.219 – 0.763) | 0.01 | 0.991  (0.796 – 1.235) | 0.90 |

### 8.)  Probability of events

#### Table S14. The 2-, 5-, 7-, and 10-year event probabilities and corresponding 95% confidence intervals (CIs) for all-cause mortality, reoperation, myocardial infarction, stroke, and heart failure

|  |  |  | Years | | | |
| --- | --- | --- | --- | --- | --- | --- |
| Event | Group |  | 2 | 5 | 7 | 10 |
| All-cause Mortality | sB-ARV | Number at Risk | 9789 | 5810 | 3551 | 969 |
|  |  | Event Probability | 0.147 | 0.319 | 0.458 | 0.695 |
|  |  | 95% CI | 0.136 - 0.157 | 0.303- 0.334 | 0.440 - 0.475 | 0.673 - 0.716 |
|  | TAVR | Number at Risk | 2829 | 878 | 315 | 37 |
|  |  | Event Probability | 0.194 | 0.459 | 0.631 | 0.842 |
|  |  | 95% CI | 0.181 - 0.206 | 0.440 - 0.478 | 0.606 - 0.655 | 0.807- 0.871 |
| Reoperation | sB-ARV | Number at Risk | 8738 | 4780 | 2639 | 423 |
|  |  | Event Probability | 0.010 | 0.0145 | 0.022 | 0.039 |
|  |  | 95% CI | 0.008 - 0.011 | 0.012 - 0.017 | 0.019 - 0.026 | 0.033 - 0.045 |
|  | TAVR | Number at Risk | 3608 | 1005 | 319 | 16 |
|  |  | Event Probability | 0.007 | 0.010 | 0.010 | 0.011 |
|  |  | 95% CI | 0.005 - 0.009 | 0.007 - 0.013 | 0.007 - 0.013 | 0.008 - 0.016 |
| Myocardial Infarction | sB-ARV | Number at Risk | 9327 | 5304 | 3208 | 750 |
|  |  | Event Probability | 0.013 | 0.022 | 0.029 | 0.036 |
|  |  | 95% CI | 0.011 - 0.015 | 0.020 - 0.025 | 0.025 - 0.032 | 0.031 - 0.040 |
|  | TAVR | Number at Risk | 4252 | 1250 | 415 | 34 |
|  |  | Event Probability | 0.011 | 0.024 | 0.030 | 0.033 |
|  |  | 95% CI | 0.009 - 0.014 | 0.020 - 0.029 | 0.024 - 0.036 | 0.027 - 0.041 |
| Stroke | sB-ARV | Number at Risk | 9059 | 4984 | 2968 | 681 |
|  |  | Event Probability | 0.041 | 0.082 | 0.102 | 0.125 |
|  |  | 95% CI | 0.038 - 0.045 | 0.077 - 0.088 | 0.096 - 0.108 | 0.117 - 0.133 |
|  | TAVR | Number at Risk | 4097 | 1165 | 388 | 31 |
|  |  | Event Probability | 0.052 | 0.093 | 0.103 | 0.111 |
|  |  | 95% CI | 0.046 - 0.057 | 0.085 - 0.101 | 0.094 - 0.112 | 0.100 - 0.122 |
| Heart failure | sB-ARV | Number at Risk | 7208 | 3925 | 2262 | 555 |
|  |  | Event Probability | 0.141 | 0.202 | 0.255 | 0.312 |
|  |  | 95% CI | 0.134 - 0.148 | 0.193 - 0.210 | 0.245 - 0.264 | 0.300 - 0.324 |
|  | TAVR | Number at Risk | 2815 | 759 | 244 | 18 |
|  |  | Event Probability | 0.177 | 0.288 | 0.337 | 0.380 |
|  |  | 95% CI | 0.167 - 0.188 | 0.273 - 0.303 | 0.319 - 0.355 | 0.355 - 0.405 |

#### Table S15. The 2-, 5-, 7-, and 10-year all-cause mortality probabilities and corresponding 95% confidence intervals (CIs) within the different subgroups; NA (“not available”) indicates that the corresponding all-cause mortality probability could not be calculated because of a too-short follow-up

| All-cause Mortality | | | | | | |
| --- | --- | --- | --- | --- | --- | --- |
|  |  |  | Years | | | |
| Subgroup | Group |  | 2 | 5 | 7 | 10 |
| Patients 2010 - 2015 | sB-ARV | Number at Risk | 4892 | 4057 | 3431 | 969 |
|  |  | Event Probability | 0.127 | 0.276 | 0.388 | 0.579 |
|  |  | 95% CI | 0.119 - 0.136 | 0.265 - 0.288 | 0.375 - 0.401 | 0.564 - 0.594 |
|  | TAVR | Number at Risk | 1212 | 750 | 484 | 51 |
|  |  | Event Probability | 0.242 | 0.531 | 0.697 | 0.880 |
|  |  | 95% CI | 0.221 - 0.263 | 0.506 - 0.555 | 0.674 - 0.719 | 0.856 - 0.899 |
| Patients 2015 - 2020 | sB-ARV | Number at Risk | 4897 | 1753 | 120 | NA |
|  |  | Event Probability | 0.106 | 0.227 | 0.345 | NA |
|  |  | 95% CI | 0.098 - 0.114 | 0.214 - 0.240 | 0.320 - 0.368 | NA |
|  | TAVR | Number at Risk | 3559 | 722 | 30 | NA |
|  |  | Event Probability | 0.211 | 0.478 | 0.680 | NA |
|  |  | 95% CI | 0.200 - 0.222 | 0.459 - 0.496 | 0.641 - 0.716 | NA |
| Patients with Diabetes | sB-ARV | Number at Risk | 1690 | 941 | 564 | 122 |
|  |  | Event Probability | 0.157 | 0.336 | 0.474 | 0.685 |
|  |  | 95% CI | 0.141 - 0.172 | 0.314 - 0.357 | 0.448 - 0.499 | 0.651 - 0.715 |
|  | TAVR | Number at Risk | 914 | 238 | 84 | 6 |
|  |  | Event Probability | 0.251 | 0.591 | 0.744 | 0.924 |
|  |  | 95% CI | 0.228 - 0.274 | 0.556 - 0.622 | 0.706 - 0.778 | 0.868 - 0.956 |
| Patients with pulmonary disease | sB-ARV | Number at Risk | 275 | 66 | 38 | NA |
|  |  | Event Probability | 0.133 | 0.317 | 0.488 | NA |
|  |  | 95% CI | 0.096 - 0.168 | 0.252 - 0.376 | 0.391 - 0.569 | NA |
|  | TAVR | Number at Risk | 257 | 23 | 11 | NA |
|  |  | Event Probability | 0.309 | 0.602 | 0.714 | NA |
|  |  | 95% CI | 0.265 - 0.351 | 0.513 - 0.675 | 0.600 - 0.795 | NA |
| Patients with kidney diseases | sB-ARV | Number at Risk | 894 | 390 | 218 | 46 |
|  |  | Event Probability | 0.217 | 0.426 | 0.556 | 0.754 |
|  |  | 95% CI | 0.193 - 0.240 | 0.394 - 0.456 | 0.519 - 0.590 | 0.706 - 0.793 |
|  | TAVR | Number at Risk | 893 | 222 | 67 | 6 |
|  |  | Event Probability | 0.306 | 0.618 | 0.770 | 0.915 |
|  |  | 95% CI | 0.282 - 0.330 | 0.585 - 0.649 | 0.731 - 0.804 | 0.857 - 0.949 |
